# Adult disability and mental/psychological illness in Somaliland: socioeconomic inequalities and co-occurrence in the 2020 Somaliland Health and Demographic Survey

**DOI:** 10.64898/2026.09.24.26363961

**Authors:** Ahmed Adan Saed, Omer Abdulkadir Osman, Abdimalik Mohamed Adan, Yousuf Ismael Suleiman, Ahmed Ibrahim Mohamed

## Abstract

Disability is an important population-health concern, but adult-specific evidence on its distribution and socioeconomic patterning remains limited in Somaliland. This study estimated the prevalence and distribution of reported disability among adults in Somaliland and examined socioeconomic inequality, reported mental/psychological illness, and their co-occurrence.

We conducted a cross-sectional secondary analysis of the 2020 Somaliland Health and Demographic Survey. The analysis included 14,579 adults aged ≥18 years who were usual residents, had the disability module administered, and had a defined reported mental/psychological illness measure. Reported disability was defined as at least one positive response across seven disability domains: sight, hearing, speech, learning, mobility, self-care, and mental. Survey-weighted prevalence estimates and design-based comparisons were calculated. Survey-weighted logistic regression assessed associations with household wealth and other covariates. Socioeconomic inequality was assessed using a concentration index and the normalized Erreygers concentration index. Co-occurrence was evaluated by comparing observed joint prevalence with that expected under statistical independence and by survey-weighted logistic regression.

The survey-weighted prevalence of any reported disability was 7.30% (95% CI 6.65–8.02). Sight was the most frequently reported domain (3.83%, 95% CI 3.19–4.59), followed by hearing (1.54%, 95% CI 1.20–1.97) and mobility (1.39%, 95% CI 1.14–1.70). Disability prevalence increased markedly with age, from 4.15% among adults aged 18–29 years to 23.96% among those aged ≥60 years. The crude wealth pattern was non-monotonic and was not statistically detectable after adjustment for residence and region. The concentration index for reported disability was 0.0584 (SE 0.0284, p=0.0415). Reported mental/psychological illness had a weighted prevalence of 0.71% (95% CI 0.50–1.00). Both reported disability and reported mental/psychological illness occurred in 0.5032% of adults, compared with 0.05185% expected under statistical independence; the observed-to-expected ratio was 9.70 (95% CI 7.60–11.81). After adjustment for age group and sex, reported disability was strongly associated with reported mental/psychological illness (OR 42.99, 95% CI 18.45–100.18).

Reported disability affected approximately one in fourteen adults in Somaliland and showed a marked age gradient and substantial geographic variation. The crude socioeconomic pattern was attenuated after adjustment for geographic context, while concentration analysis indicated modest marginal concentration toward higher wealth ranks. Reported mental/psychological illness was uncommon but co-occurred with reported disability more often than expected under independence. These findings provide an adult population baseline for Somaliland while highlighting the need to distinguish survey-reported disability from the separate mental/psychological illness construct and to interpret the findings within the limitations of cross-sectional, survey-reported measures.

**Author Summary:** Why was this study done?

- Evidence on disability among adults in Somaliland is limited, especially on its socioeconomic distribution and relationship with reported mental/psychological illness.
- We used the 2020 Somaliland Health and Demographic Survey to examine these patterns nationally.

What did the researchers do and find?

- We analysed 14,579 adults aged 18 years or older using survey-weighted methods that accounted for the survey’s sampling design.
- Overall, 7.30% of adults reported at least one disability domain. Sight, hearing, and mobility were most frequently reported, and disability was much more common among older adults and varied across regions and residence settings.
- Reported mental/psychological illness was found in 0.71% of adults. Both conditions occurred together in 0.50%, substantially more than expected under statistical independence.

What do these findings mean*?*

- Our study provides an adult population baseline for reported disability in Somaliland and shows clear differences by age and geographic context.
- Our findings highlight the importance of keeping reported disability and reported mental/psychological illness as distinct measures.
- Because our data are cross-sectional and based on reported survey measures, the findings describe the 2020 survey period and do not establish clinical diagnoses or causal relationships.

## Introduction

Disability is a major population-health concern and is associated with poorer health, greater functional limitation, and substantial inequities in access to health and social services. The World Health Organization estimates that approximately 1.3 billion people, or 16% of the global population, experience significant disability. These inequities arise in part from stigma and discrimination, poverty and social exclusion, exclusion from education and employment, inaccessible environments, and barriers within health systems (1,2).

Estimating the prevalence of disability is therefore only one component of understanding its population burden. Its social and economic distribution is also important because disability may be patterned by household socioeconomic position, educational attainment, place of residence, and geographic context. Evidence from ten sub-Saharan African household surveys found that disability was more common in rural populations and among people with lower educational attainment and increased sharply with age, while reported disability rates were broadly similar across poorer and wealthier households (3). These findings illustrate why the socioeconomic distribution of disability should be estimated empirically rather than assumed. For binary health outcomes, concentration curves and concentration indices provide established approaches for quantifying socioeconomic-related inequality; corrected forms of the concentration index have also been proposed for bounded health variables (3,4).

The relationship between disability and mental/psychological illness warrants separate consideration. These constructs are not necessarily equivalent: a disability measure may capture functional difficulty or limitation, whereas mental/psychological illness represents a health-condition construct. Nevertheless, international population-based evidence has demonstrated a substantial association between mental and physical disorders and disability. In the World Mental Health Surveys, adults with mental–physical co-morbidity had greater disability than those with individual conditions alone, although the magnitude and form of the association varied according to the disorders and statistical model considered (5).

Evidence concerning disability in Somalia and Somaliland remains comparatively limited and uses heterogeneous populations and data sources. A 2026 analysis of the 2022 Somali Integrated Household Budget Survey estimated disability among 19,832 adults aged 18 years or older in Somalia, demonstrating that nationally representative adult disability research is feasible in the Somali context but using data that are distinct from the Somaliland survey examined in the present study (6). In Somaliland, a 2026 descriptive study of 44 persons with disabilities in Hargeisa examined socioeconomic exclusion and barriers to education, employment, healthcare access, and social participation, but its small, localized sample cannot provide a population-level estimate for Somaliland (7).

Mental-health evidence from the Somali and Somaliland context is similarly heterogeneous. A 2020 publication describing mental health in Somaliland highlighted substantial unmet mental-health needs but was not a nationally representative prevalence study (8). More recently, a national analysis of the 2020 Somali Health and Demographic Survey reported self-reported mental-health difficulties among people aged 10 years and older, again using the Somali survey rather than the Somaliland survey and a different outcome definition (9). These differences in population, setting, survey instrument, and outcome definition make direct numerical comparisons with the present study inappropriate.

The Somaliland Health and Demographic Survey (SLHDS) 2020 provides an important national source for addressing the evidence gap. The survey was conducted by the Government of Somaliland through the Central Statistics Department of the Ministry of Planning and National Development and the Ministry of Health Development, with UNFPA and cooperating partners. It was the first Demographic and Health Survey-type survey conducted in Somaliland and covered six regions and three residence domains: urban, rural, and nomadic (10).

The SLHDS 2020 report already contains all-age descriptive information on disability, including disability domains and distributions by selected demographic and socioeconomic characteristics, as well as information on reported disability origin, age at onset, and care or support (10). The survey records seven disability domains—sight, hearing, speech, learning, mobility, self-care, and mental—and also records a separate mental/psychological illness category within its chronic-disease module(10). The two constructs are therefore distinguishable within the source data and should not be treated as interchangeable.

What has remained insufficiently addressed is the adult-focused analytical treatment of these data. The published SLHDS findings do not provide an adult-specific survey-weighted analysis integrating prevalence, domain distribution, socioeconomic patterning, concentration-based inequality, and the relationship between reported disability and the separately recorded mental/psychological illness construct. In particular, the joint occurrence of these two constructs and whether their observed co-occurrence exceeds that expected under statistical independence have not previously been examined in an adult Somaliland population using SLHDS 2020. Based on the literature search conducted for this study through September 2026, no identified publication directly combines these components in Somaliland (6,7,9,10).

The distinction between reported disability and reported mental/psychological illness is central to the present analysis. The SLHDS disability module records the seven disability domains, including a separate mental domain, whereas mental/psychological illness is recorded as a distinct condition within the chronic-disease module (10). The present study consequently treats the mental disability domain and reported mental/psychological illness as separate constructs and examines their co-occurrence without assuming a temporal or causal direction. This distinction is important because the cross-sectional survey design cannot establish whether one condition preceded or caused the other.

Accordingly, this study aimed to estimate the prevalence of reported disability among adults aged 18 years and older in Somaliland; describe the distribution of the seven reported disability domains by sex and age; examine the socioeconomic distribution of reported disability across household wealth, education, residence, and region; quantify socioeconomic inequality using concentration-based measures; estimate the prevalence of reported mental/psychological illness; and examine the co-occurrence of reported disability and reported mental/psychological illness, including whether their observed joint occurrence exceeds that expected under statistical independence and whether the co-occurring group varies across socioeconomic and contextual characteristics. The study was conducted as a cross-sectional secondary analysis of SLHDS 2020 and was designed to describe associations rather than establish causality.

## Methods

### Study design and data source

We conducted a cross-sectional secondary analysis of the 2020 Somaliland Health and Demographic Survey (SLHDS 2020), a nationally representative household survey conducted in Somaliland. The SLHDS used a stratified multistage probability cluster design covering the six regions of Somaliland and three residence domains—urban, rural, and nomadic. The SLHDS main survey data collection in urban and rural areas was conducted from August 2018 to December 2019, with nomadic data collection conducted as part of the corresponding main-survey fieldwork (10). The disability and chronic-disease information used in this study was collected through the household questionnaire on household members rather than through a clinical examination (10).

The present analysis was restricted to adults aged 18 years or older and was designed to estimate reported disability and its socioeconomic distribution, while examining reported mental/psychological illness as a separate construct and its co-occurrence with reported disability. Because the source survey is cross-sectional, all analyses were interpreted as associations or distributions rather than causal effects.

### Study population and analytic sample

The source person-level file contained 35,381 household-member records. Records with unusable age information were excluded, after which adults aged 18 years or older were identified. The eligibility rule for the present analysis required that an individual was aged ≥18 years, was a usual resident (HV102=1), had the disability module administered, and had an answered chronic-disease gate such that the reported mental/psychological illness measure could be defined.

The resulting analytic sample comprised 14,579 adults. Of the 15,041 records aged ≥18 years with usable age information, 342 were excluded because they were not usual residents and six because the disability module was not administered; a further 114 records with a chronic-disease gate coded as “Don’t know” were excluded because the mental/psychological illness variable could not be defined consistently. The final denominator therefore consisted of adults aged ≥18 years who were usual residents, had the disability module administered, and had a defined mental/psychological illness measure. Because this was a secondary analysis of an existing population-based survey, the analytic sample comprised all eligible records available in the source dataset; no a priori sample-size or power calculation was performed.

We used the usual-resident (de jure) definition rather than the de facto household population because disability was treated as a stable personal characteristic rather than a one-night household exposure. The de facto adult count of 14,317 was retained as a reference during data preparation, but a de facto sensitivity analysis was not possible from the frozen final analytic file because excluded records were not retained in that file.

### Survey design, weights, and complex-sample estimation

All estimates incorporated the SLHDS sampling design using sampling weights, stratification, and clustering. The final analytic dataset contained 18 sampling strata, corresponding to the six regions crossed by the three residence domains, and the survey-weighted analyses used 188 stratum-specific PSU units. The final analytic dataset contained 18 sampling strata and 188 unique stratum-specific PSU units. The raw cluster identifier (HV001) was not unique across sampling strata; therefore, PSU units in the analysis were represented at the stratum-specific level.

The survey declaration used the final analysis variables survey_weight, survey_psu, and survey_strata, with single-unit(centered) specified for strata containing a single sampled PSU. The same survey declaration was retained throughout the analysis.

### Definition of reported disability

Reported disability was the primary outcome. It was constructed from the seven labelled disability-domain indicators corresponding to the SLHDS disability items: sight, hearing, speech, learning, mobility, self-care, and mental. An individual was classified as having any reported disability when at least one of these seven domain indicators was positive. Individuals with the disability module administered but no positive domain were coded as having no reported disability. No severity threshold was imposed because the audited SLHDS materials did not provide a graded severity scale or establish the use of the Washington Group Short Set(10).

The labelled per-domain indicators were treated as authoritative because the concatenated HV408 multi-response string was stored in a width-limited field and could truncate multiple selections. Consequently, the derived disability variables were based on the labelled domain indicators rather than the concatenated string.

Seven domain-specific binary outcomes were retained for secondary analyses. The seven domains were not treated as mutually exclusive, because an individual could report more than one domain. A count of the number of reported disability domains was retained as a descriptive variable only and was not interpreted as a severity measure.

An internal source-data inconsistency was identified in which the SLHDS “None” response coexisted with at least one positive disability-domain indicator in 227 adults. Following the predefined data-preparation rule, the positive domain indicators were treated as the primary disability information, while the None indicator was retained for quality-control purposes. A sensitivity analysis excluding these 227 records was subsequently performed.

### Definition of reported mental/psychological illness

Reported mental/psychological illness was treated as a separate secondary outcome and was not derived from the mental disability domain. It was constructed from the chronic-disease condition indicator HV402Q, labelled “Mental/Psychological illness,” among individuals for whom the chronic-disease gate HV401 indicated that they were suffering from a chronic disease. Thus, an individual was coded positive when HV401=1 and HV402Q=1; a negative value was assigned when the chronic-disease gate was positive but HV402Q was not selected, or when HV401=2. Records with an unresolved “Don’t know” chronic-disease response were excluded from the final analytic population.

The separate physician-information variable HV403 was not used to define this outcome. Accordingly, the manuscript refers to this construct as reported mental/psychological illness, rather than physician-diagnosed mental illness.

### Disability–mental/psychological illness co-occurrence

The two constructs were cross-classified into four mutually exclusive categories:

1. Neither: no reported disability and no reported mental/psychological illness;
2. Disability only: reported disability without reported mental/psychological illness;
3. Mental/psychological illness only: reported mental/psychological illness without reported disability; and
4. Both: reported disability and reported mental/psychological illness.

The constructs were retained as distinct measures throughout the analysis and were not combined into a single clinical diagnosis.

### Socioeconomic and demographic variables

The primary socioeconomic exposure was household wealth index quintile. The SLHDS wealth index is an asset-based relative measure rather than a direct measure of income or consumption. Wealth quintile was used as the principal categorical socioeconomic variable, while the continuous wealth-index factor score was used as the ranking variable for concentration-based inequality analysis(10).

Secondary socioeconomic variables were educational attainment, residence, and region. Education was classified as no education, primary, secondary, or higher education; the non-formal “Koranic” category was retained as missing rather than incorporated into formal schooling categories. Residence was categorized as urban, rural, or nomadic, and region was categorized as Awdal, Marodijeh, Sahil, Togdheer, Sool, or Sanaag.

Demographic and social covariates included age, sex, marital status, household size, and relationship to the household head. Age was examined both continuously and in four ordered categories: 18-29, 30-44, 45-59, and ≥60 years. The primary multivariable disability model included wealth, age group, sex, education, residence, and region. Marital status, household size, and relationship to the household head were used in prespecified sensitivity specifications rather than as mandatory components of the primary model.

### Contextual disability variables

Among adults reporting disability, the analysis additionally described three contextual characteristics recorded in the SLHDS disability module: reported origin of disability, age at onset, and care/support received. The origin categories followed the source coding and included congenital, contagious disease, child-birth conditions, other disease, abuse, aging, injury/accident, witchcraft, other, and don’t know.

Age at onset was grouped using categories consistent with the SLHDS reporting structure: <5, 5– 9, 10-19, 20-29, 30-39, 40-49, 50-59, 60-69, and ≥70 years. The source survey asks disabled household members about their age when the disability began(10).

Care/support variables covered medical, welfare, financial, nutritional, and no-support responses. These were treated as multi-response indicators, rather than as mutually exclusive categories, because multiple types of support could be recorded for the same individual.

### Statistical analysis

All analyses were conducted using Stata/MP version 17.0 (StataCorp, College Station, TX, USA), and the Stata analysis code used in this is publicly available in Zenodo https://doi.org/10.5281/zenodo.22906585. All analysis incorporated the complex survey design through the svy framework with linearized variance estimation. Estimates are presented with survey-adjusted 95% confidence intervals unless otherwise specified. All hypothesis tests were two-sided, and p<0.05 was considered statistically significant. No formal adjustment for multiple comparisons was applied; domain-specific, sex- and age-stratified, and other secondary analyses were interpreted as exploratory, and their p values were not treated as evidence from independent confirmatory hypotheses.

### Descriptive analysis

We first described the analytic population according to age, sex, educational attainment, household wealth quintile, residence, and region. Survey-weighted proportions were then estimated for:

- any reported disability;
- each of the seven disability domains;
- reported mental/psychological illness; and
- the four co-occurrence categories.

Domain-specific disability prevalence was examined separately by sex and age group, with design-based tests of differences between groups. The four ordered age groups were additionally used to assess the prespecified age gradient for the primary disability outcome.

### Socioeconomic distribution and multivariable analysis of disability

Reported disability prevalence was estimated across wealth quintiles, education categories, residence domains, and regions. Survey-weighted logistic regression was then used to examine associations between reported disability and the socioeconomic and demographic covariates.

The primary multivariable model included household wealth quintile, age group, sex, education, residence, and region simultaneously. Alternative specifications examined the wealth association under different adjustment sets, including models additionally or separately incorporating education, residence, and region. Adjusted predicted probabilities across wealth quintiles were estimated from the primary model.

A separate sensitivity specification additionally adjusted for marital status and household size, and a further specification included relationship to the household head. These models were intended to assess the robustness of the estimated wealth association to the inclusion of additional social and household-context covariates rather than to establish causal pathways.

### Socioeconomic inequality analysis

Socioeconomic inequality in reported disability was quantified using a concentration index, with the continuous wealth-index factor score as the ranking variable. The concentration curve was plotted against the cumulative population ranked from lower to higher wealth. Because reported disability is a bounded binary outcome, the normalized Erreygers concentration index was also estimated (4). Standard errors and statistical inference for the concentration indices were estimated using the survey design specified for the analysis, incorporating the sampling weights, stratification and clustering.

The concentration analysis was interpreted together with the direction of the concentration curve and the sign of the index. No categorical wealth-quintile gradient was assumed in advance because concentration-based inequality and categorical prevalence patterns address related but distinct aspects of socioeconomic distribution.

### Mental/psychological illness analysis

The prevalence of reported mental/psychological illness was estimated overall and according to age, sex, education, residence, region, and household wealth. Survey-weighted logistic regression models examined its association with socioeconomic and demographic variables.

Because the mental/psychological illness outcome was uncommon, estimates were interpreted with attention to the relatively small number of positive observations and correspondingly wide confidence intervals.

### Co-occurrence and independence analysis

The four-category co-occurrence outcome was examined across wealth, education, residence, and region using survey-weighted descriptive comparisons and design-based tests of association. A survey-weighted multinomial logistic model was used for the categorical wealth analysis.

To test the prespecified expectation that disability and reported mental/psychological illness would co-occur more frequently than expected under statistical independence, we compared the observed joint prevalence with the prevalence expected under independence, calculated as the product of the two marginal survey-weighted prevalences. The observed-to-expected ratio was estimated as the ratio of the observed joint prevalence to the product of the two marginal survey-weighted prevalences. The ratio and its 95% confidence interval were estimated using Stata’s nonlinear combination (nlcom) procedure, with uncertainty propagated from the estimated marginal and joint prevalences.

An adjusted survey-weighted logistic model then examined the association between reported disability and reported mental/psychological illness while adjusting for age group and sex. This model was interpreted as an association model, not a causal model, because the cross-sectional design provides no temporal ordering. A secondary sensitivity model additionally adjusted for marital status and household size.

### Contextual and sensitivity analyses

Among adults with reported disability and available contextual information, survey-weighted distributions were estimated for reported disability origin, age at onset, and support received.

Sensitivity analysis excluding the 227 records in which the source “None” indicator co-occurred with a positive disability-domain indicator was undertaken to evaluate the effect of this internal source-data inconsistency on the primary disability estimate and its socioeconomic association.

The final frozen analytic dataset did not retain enough information to reconstruct the previously identified de facto-residency, chronic-disease gate-lifting, or alternative Koranic-education coding sensitivity analyses. These were therefore not represented as completed analyses.

### Missing data

No statistical imputation was performed. The core analysis variables—including reported disability, reported mental/psychological illness, co-occurrence, survey weight, PSU, and stratum—were complete in the final analytic dataset. Missingness was concentrated in education and contextual disability variables, the latter of which were only applicable to respondents reporting disability. Education contained 180 missing records (approximately 1.2% of the analytic sample), largely corresponding to the “Don’t know” coding retained as missing during data preparation.

The primary multivariable analysis therefore used complete observations for the variables required by each model, and no missing-value imputation was introduced.

### Ethics Statement

This study is a secondary analysis of anonymized data from the 2020 Somaliland Health and Demographic Survey (SLHDS). Ethical approval for the original SLHDS was obtained by the Central Statistics Department (CSD), Ministry of Planning and National Development, in collaboration with the Ministry of Health Development (MoHD), Republic of Somaliland, prior to data collection. Permission to access and analyse the anonymized SLHDS dataset for this study was obtained from the Central Statistics Department. Because the present study involved secondary analysis of fully de-identified data, no additional ethical approval or informed consent from participants was required.

## Results

### Analytic population

The final analytic sample comprised 14,579 adults aged 18 years or older. Of the 35,381 person records in the source person-level file, 15,041 were aged ≥18 years and 14,699 were usual residents. Six adults were excluded because the disability module was not administered, and 114 were excluded because the chronic-disease screening gate was coded as “Don’t know,” leaving 14,579 adults for the final analysis. The participant selection and analytical sample flow are shown in Fig 1. The analytic design represented 18 sampling strata and 188 stratum-specific primary sampling-unit units.

**Fig 1.**
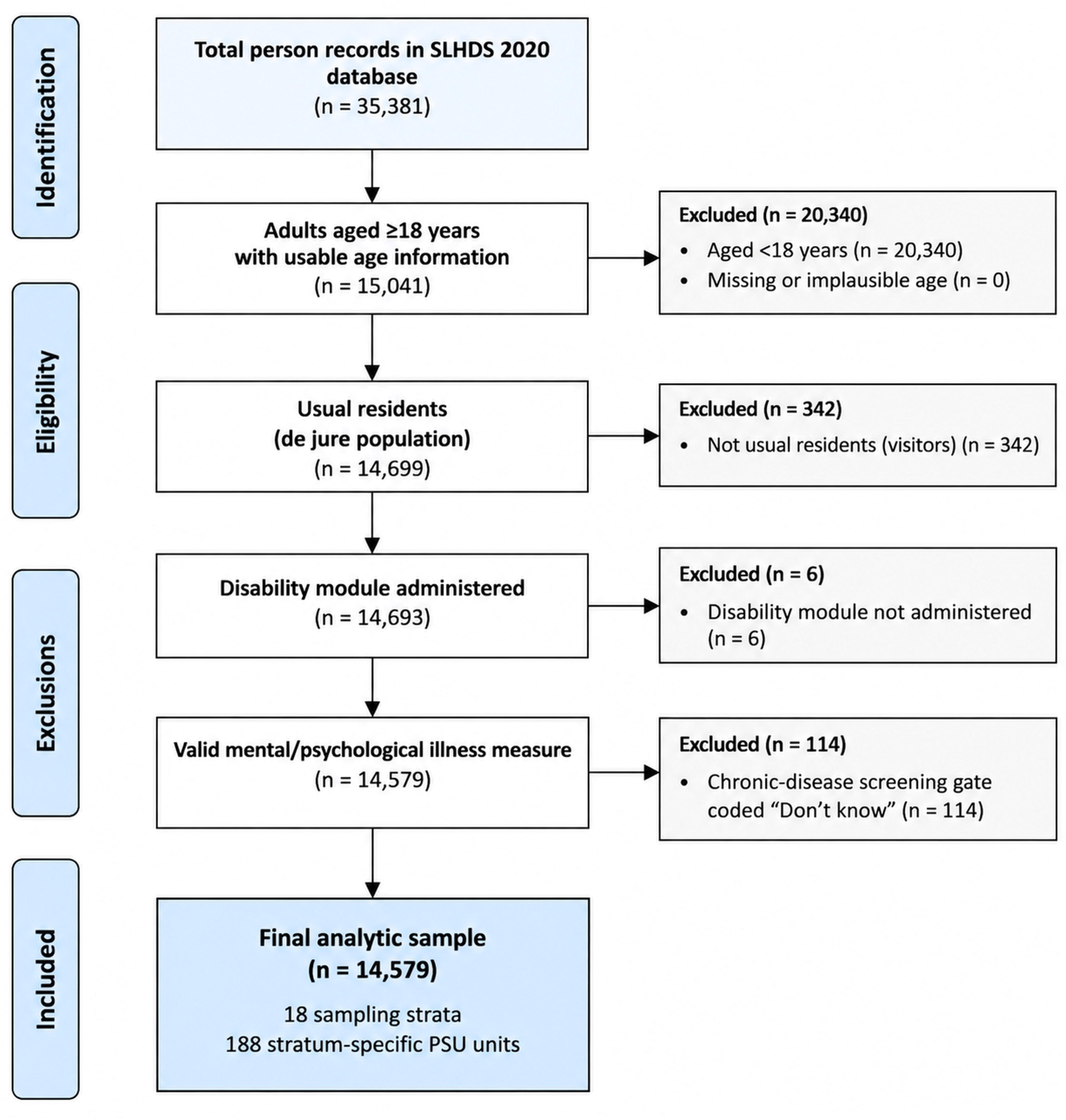
participant-selection and analytical flow diagram.

Men accounted for 46.01% of the sample and women for 53.99%. Adults aged 18–29 years comprised 39.46%, those aged 30–44 years 30.15%, those aged 45–59 years 16.31%, and those aged ≥60 years 14.08%. Most adults had no formal education (74.71%). Residence was distributed across urban (36.91%), rural (30.66%) and nomadic (32.43%) populations. The six regions contributed between 12.30% and 21.54% of the sample, and education was missing for 180 adults (1.23%). The characteristics of the analytic sample are presented in Table 1.

**Table 1.** Characteristics of the adult analytic sample, SLHDS 2020.

| Characteristic | Category | N | % |
| --- | --- | --- | --- |
| Sex | Male | 6,708 | 46.01 |
|  | Female | 7,871 | 53.99 |
| Age group (years) | 18–29 | 5,753 | 39.46 |
|  | 30–44 | 4,396 | 30.15 |
|  | 45–59 | 2,378 | 16.31 |
| | $\geq 60$ | 2,052 | 14.08 |
| Education | No education | 10,892 | 74.71 |
|  | Primary | 1,588 | 10.89 |
|  | Secondary | 1,199 | 8.22 |
|  | Higher | 720 | 4.94 |
|  | Missing | 180 | 1.23 |
| Household wealth quintile | Lowest | 6,146 | 42.16 |
|  | Second | 1,521 | 10.43 |
|  | Middle | 1,839 | 12.61 |
|  | Fourth | 2,481 | 17.02 |
|  | Highest | 2,592 | 17.78 |
| Residence | Urban | 5,381 | 36.91 |
|  | Rural | 4,470 | 30.66 |
|  | Nomadic | 4,728 | 32.43 |
| Region | Awdal | 2,196 | 15.06 |
|  | Marodijeh | 2,036 | 13.97 |
|  | Sahil | 1,793 | 12.30 |
|  | Togdheer | 2,442 | 16.75 |
|  | Sool | 2,971 | 20.38 |
|  | Sanaag | 3,141 | 21.54 |

**Note:** *n* and percentages are unweighted sample counts and proportions. The analytic sample comprised 14,579 adults; education was missing for 180 participants.

### Prevalence and composition of reported disability

The survey-weighted prevalence of any reported disability was 7.30% (95% CI 6.65–8.02). Sight was the most frequently reported disability domain, affecting 3.83% (95% CI 3.19–4.59), followed by hearing at 1.54% (95% CI 1.20–1.97) and mobility at 1.39% (95% CI 1.14–1.70). Speech, learning and self-care difficulties were less frequently reported, while the separate mental disability domain was reported by 1.29% of adults (95% CI 0.98–1.69). The disability domains were not mutually exclusive. These estimates are presented in Table 2.

**Table 2.** Survey-weighted prevalence of reported disability, reported mental/psychological illness, and co-occurrence among adults.

| Outcome | Unweighted n | Weighted prevalence (%) | 95% CI |
| --- | --- | --- | --- |
| Any reported disability | 947 | 7.30 | 6.65–8.02 |
| Sight disability | 479 | 3.83 | 3.19–4.59 |
| Hearing disability | 217 | 1.54 | 1.20–1.97 |
| Speech disability | 49 | 0.36 | 0.23–0.56 |
| Learning disability | 44 | 0.35 | 0.22–0.56 |
| Mobility disability | 246 | 1.39 | 1.14–1.70 |
| Self-care disability | 71 | 0.29 | 0.21–0.40 |
| Mental disability domain | 156 | 1.29 | 0.98–1.69 |
| Reported mental/psychological illness | 99 | 0.71 | 0.50–1.00 |
| Neither | 13,601 | 92.49 | 91.74–93.18 |
| Disability only | 879 | 6.80 | 6.16–7.50 |
| Mental/psychological illness only | 31 | 0.21 | 0.12–0.34 |
| Both conditions | 68 | 0.50 | 0.32–0.78 |

**Note:** Estimates incorporate survey weights, stratification and clustering. The seven disability-domain indicators are not mutually exclusive. The four co-occurrence categories are mutually exclusive and exhaustive. Reported mental/psychological illness refers to the separate chronic-disease item and is distinct from the mental disability domain.

Reported mental/psychological illness, measured separately through the chronic-disease module, had a weighted prevalence of 0.71% (95% CI 0.50–1.00). In the four-category joint classification, 92.49% of adults had neither reported disability nor reported mental/psychological illness, 6.80% had disability only, 0.21% had reported mental/psychological illness only, and 0.50% had both conditions (Table 2).

### Age pattern

Weighted prevalence of any reported disability increased from 4.15% among adults aged 18–29 years to 4.67% among those aged 30–44 years, 6.43% among those aged 45–59 years, and 23.96% among adults aged ≥60 years. In the primary multivariable model, the adjusted odds ratio was 1.56 (95% CI 1.05–2.32) for adults aged 45–59 years and 6.95 (95% CI 5.05–9.56) for those aged ≥60 years compared with adults aged 18-29 years (joint *p* <0.001).

When the four age groups were entered as an ordered predictor, each one-category increase was associated with approximately twofold higher odds of reported disability (OR 1.98, 95% CI 1.78–2.21, *p*<0.001; Table S14 in S1 Table). As a sensitivity analysis, modelling age continuously yielded an OR of 1.042 per additional year (95% CI 1.035–1.049, *p*<0.001), supporting retention of categorical age in the primary model rather than imposing a linear relationship across the full age range (Table S10 in S1 Table).

### Socioeconomic and geographic distribution of reported disability

Weighted disability prevalence was 5.10% in the lowest wealth quintile, 9.10% in the second, 9.16% in the middle, 8.55% in the fourth, and 7.33% in the highest quintile. The pattern was therefore non-monotonic rather than increasing or decreasing consistently across wealth categories (Table S1 in S1 Table).

In separate univariable survey-weighted logistic regression models, the second, middle, fourth and highest wealth quintiles had higher odds of reported disability than the lowest quintile. Adults aged 45-59 years and ≥60 years also had higher odds of disability, while secondary and higher education and nomadic residence were associated with lower odds. Several regional and marital-status comparisons were statistically detectable (Table S2 in S1 Table).

Weighted disability prevalence was 8.35% in urban adults, 7.91% in rural adults and 3.57% in nomadic adults. Relative to urban adults, the univariable odds ratio was 0.94 (95% CI 0.76–1.17) for rural residence and 0.41 (95% CI 0.25–0.66) for nomadic residence. Region-specific prevalence ranged from 4.49% in Sanaag and 4.96% in Awdal **to** 8.55% in Marodijeh and 8.22% in Togdheer.

Education showed a socioeconomic pattern in crude analysis. Compared with adults with no education, the odds of reported disability were lower among adults with secondary education (OR 0.47, 95% CI 0.31–0.72) and higher education (OR 0.53, 95% CI 0.33–0.83), while the association for primary education was weaker and not conventionally statistically detectable (OR 0.71, 95% CI 0.49–1.01). The complete univariable results are provided in Table S2 in S1 Table.

### Multivariable association between household wealth and reported disability

The crude wealth pattern attenuated after adjustment for demographic and geographic characteristics. In the model adjusted for age, sex and education, the global wealth association was statistically detectable (F(4,167)=5.09, *p*=0.0007). Addition of region attenuated the association but it remained statistically detectable (F(4,167)=3.49, *p*=0.0091). When residence was included instead of region, the global wealth association was no longer statistically detectable (F(4,167)=0.53, *p*=0.7132). In the model including both region and residence, the association remained non-detectable (F(4,167)=0.63, *p*=0.6429). These alternative specifications are shown in Table 3 and Table S3 in S1 Table.

**Table 3.**
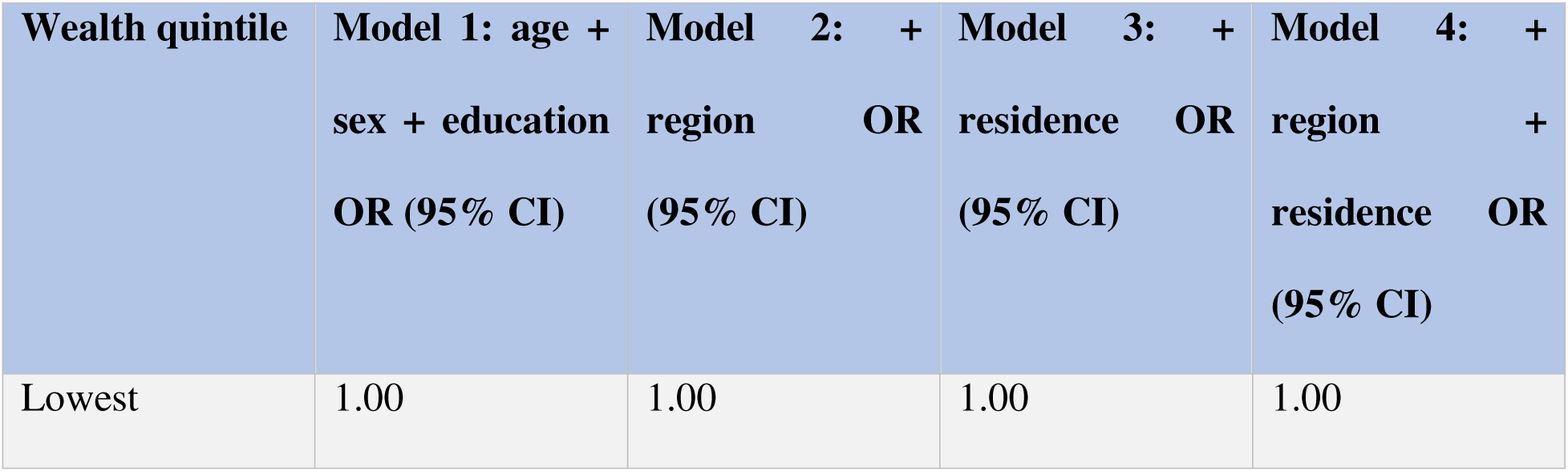
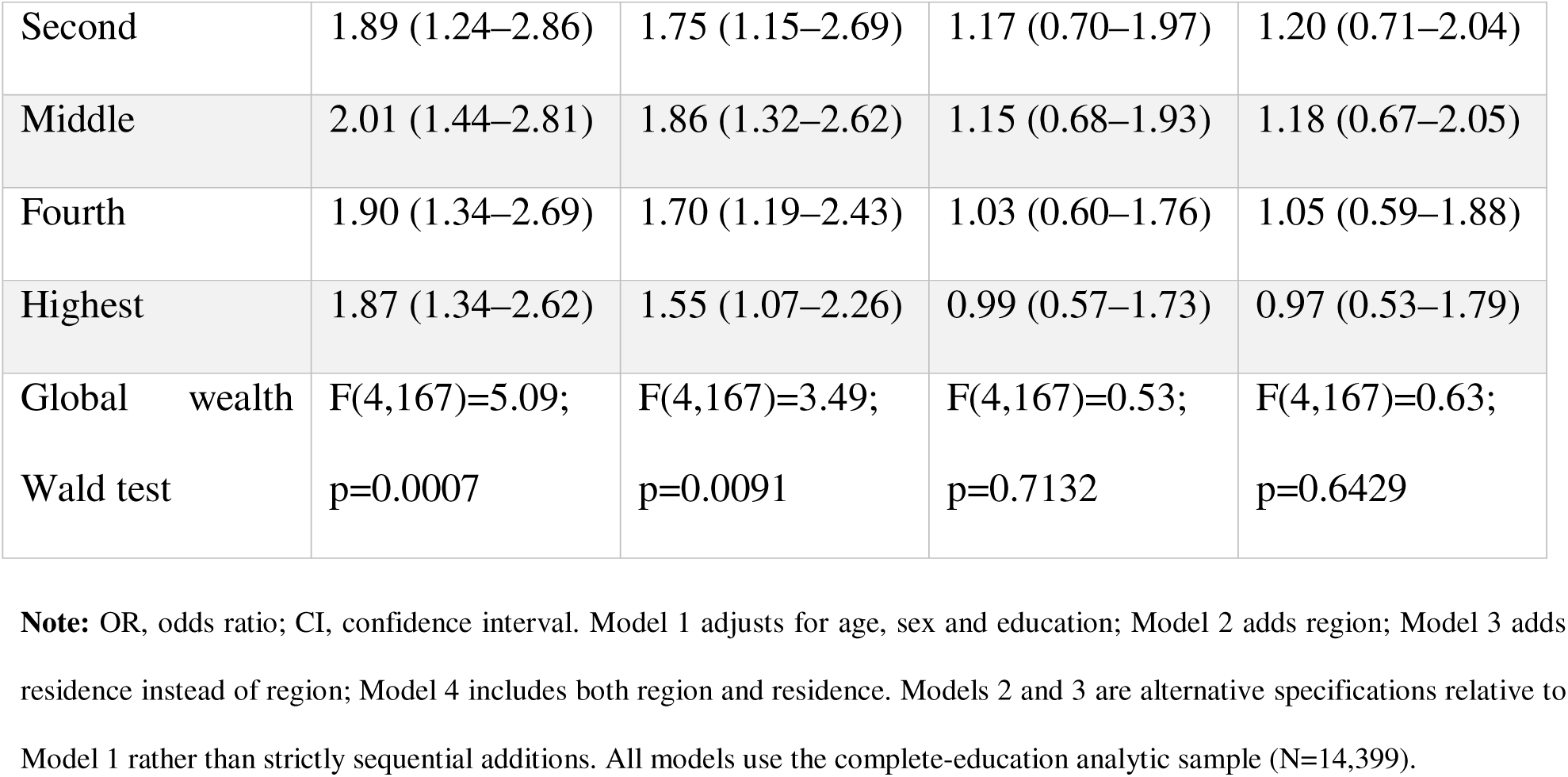
Survey-weighted logistic regression models examining the association between household wealth and reported disability under alternative adjustment sets.

The adjusted predicted probability of reported disability ranged from 7.04% in the lowest wealth quintile to 6.86% in the highest, with intermediate probabilities of 8.22%, 8.08% and 7.34% in the second, middle and fourth quintiles, respectively. No pairwise wealth contrast was statistically detectable after adjustment (Table S4 in S1 Table).

### Primary multivariable model

In the fully adjusted primary model, household wealth was not statistically associated with reported disability (global *p*=0.6429). In contrast, age, residence and region remained jointly associated with reported disability. Adults aged 45–59 years had 1.56-fold higher odds and adults aged ≥60 years had 6.95-fold higher odds compared with those aged 18-29 years. Nomadic adults had lower odds than urban adults (OR 0.42, 95% CI 0.24–0.75, *p*=0.004). Region was also jointly associated with reported disability, with higher adjusted odds in Marodijeh, Sahil, Togdheer and Sool than in Awdal. Sex and education were not statistically detectable as global predictors in the final model. The complete primary model is presented in Table 4.

**Table 4.** Primary multivariable survey-weighted logistic regression of factors associated with any reported disability.

| Predictor | Category | Adjusted OR | 95% CI | p value |
| --- | --- | --- | --- | --- |
| Household wealth quintile — global Wald test |  |  |  | 0.6429 |
|  | Lowest (reference) | 1.00 | Reference | — |
|  | Second | 1.20 | 0.71–2.04 | 0.497 |
|  | Middle | 1.18 | 0.67–2.05 | 0.568 |
|  | Fourth | 1.05 | 0.59–1.88 | 0.869 |
|  | Highest | 0.97 | 0.53–1.79 | 0.922 |
| Age group — global Wald test |  |  |  | <0.001 |
|  | 18–29 (reference) | 1.00 | Reference | — |
|  | 30–44 | 1.07 | 0.77–1.48 | 0.688 |
|  | 45–59 | 1.56 | 1.05–2.32 | 0.029 |
|  | ≥60 | 6.95 | 5.05–9.56 | <0.001 |
| Sex | Male (reference) | 1.00 | Reference | — |
|  | Female | 1.00 | 0.82–1.22 | 0.994 |
| Education —<br>global Wald test |  |  |  | 0.1068 |
|  | No education<br>(reference) | 1.00 | Reference | — |
|  | Primary | 1.02 | 0.74–1.39 | 0.916 |
|  | Secondary | 0.67 | 0.42–1.06 | 0.085 |
|  | Higher | 0.70 | 0.39–1.27 | 0.245 |
| Residence —<br>global Wald test |  |  |  | 0.0114 |
|  | Urban<br>(reference) | 1.00 | Reference | — |
|  | Rural | 0.80 | 0.60–1.07 | 0.139 |
|  | Nomadic | 0.42 | 0.24–0.75 | 0.004 |
| Region — global<br>Wald test |  |  |  | <0.001 |
|  | Awdal<br>(reference) | 1.00 | Reference | — |
|  | Marodijeh | 1.87 | 1.36–2.57 | <0.001 |
|  | Sahil | 1.73 | 1.26–2.36 | 0.001 |
|  | Togdheer | 1.84 | 1.35–2.51 | <0.001 |
|  | Sool | 1.82 | 1.32–2.52 | <0.001 |
|  | Sanaag | 1.13 | 0.81–1.58 | 0.465 |

**Note:** Primary model includes household wealth quintile, categorical age group, sex, education, residence and region. N=14,399 because education was missing for 180 adults. Global p values are survey-adjusted Wald tests. ORs describe adjusted associations and are not causal effects.

The absence of an adjusted wealth association was also robust to additional covariate specification. Adding marital status and household size produced a wealth global *p*=0.687, while additionally including relationship to household head produced *p*=0.494 (Table S21 in S1 Table).

### Concentration-based socioeconomic inequality

Although wealth was not independently associated with reported disability in the fully adjusted regression, the marginal concentration analysis indicated statistically detectable socioeconomic concentration. The concentration index for any reported disability was 0.0584 (SE 0.0284, ***p***=0.0415), with a normalized Erreygers index of 0.0171 (SE 0.0083, ***p***=0.0415). Under the study’s poorest-to-richest ranking convention, the concentration curve lay below the line of equality, consistent with relative concentration of reported disability toward the higher end of the wealth ranking. The concentration curve is shown in Figure 2, and the corresponding inequality estimates are summarized in Table S7 in S1 Table.

**Fig 2.**
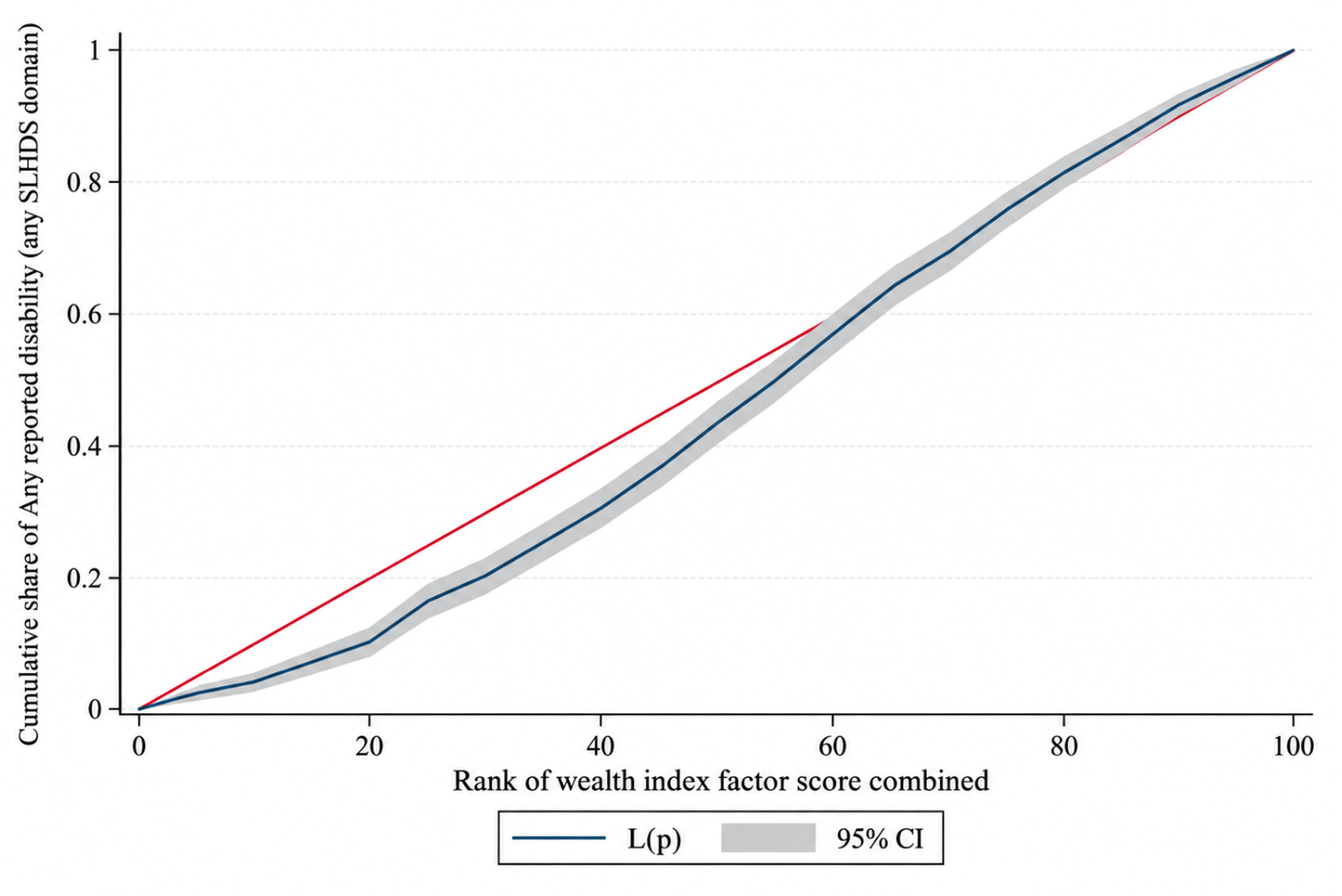
Concentration curve for any reported disability among adults aged >48 years, ranked by household wealth-index factor score. **Figure legend:** The concentration curve shows the cumulative share of reported disability against the cumulative share of adults ranked from poorest to richest according to the continuous household wealth-index factor score. The diagonal line represents equality and the shaded band the 95% confidence interval. The concentration index was 0.0584 (SE 0.0284; *p*=0.0415).

At the disability-domain level, most concentration indices were not statistically detectable. Sight, hearing, speech, learning, mobility and the mental disability domain had non-significant concentration indices. The self-care domain was the exception, with a concentration index of -0.2408 (***p***=0.0051), indicating relative concentration toward lower wealth ranks; because this was an uncommon domain, the estimate should be interpreted cautiously (Table S8 in S1 Table).

### Reported mental/psychological illness

Reported mental/psychological illness occurred in 0.71% (95% CI 0.50–1.00) of adults. The wealth-specific prevalence was 0.491% in the lowest quintile, 1.204% in the second, 1.279% in the middle, 0.780% in the fourth and 0.521% in the highest quintile (Table S1 in S1 Table).

In the multivariable analysis, reported mental/psychological illness remained uncommon and individual estimates were imprecise because only 99 events were observed. Female sex, secondary education and nomadic residence were associated with lower odds in the adjusted analyses, whereas wealth did not show a statistically detectable independent pattern. Detailed regression estimates are provided in Table S5 in S1 Table, and the distribution of the 99 events across covariate categories is shown in Table S11 in S1 Table.

The concentration index for reported mental/psychological illness was **-**0.0703 (SE 0.0685, ***p***=0.3063), with no statistically detectable socioeconomic concentration; the Erreygers-normalized index was -0.0020 (Table S7 in S1 Table).

### Co-occurrence of reported disability and reported mental/psychological illness

The observed weighted prevalence of both reported disability and reported mental/psychological illness was 0.5032%. Given marginal prevalences of 7.3045% for disability and 0.7099% for reported mental/psychological illness, the prevalence expected under statistical independence was 0.05185%. The observed-minus-expected difference was 0.45134 percentage points, and the observed-to-expected ratio was 9.70 (95% CI 7.60–11.81, *p*<0.001). A design-based Pearson test also demonstrated a statistically detectable departure from independence (F(1,170)=239.54, *p*<0.001). These results are summarized in Table 5 and detailed in Table S15 in S1 Table.

**Table 5.** Co-occurrence and association between reported disability and reported mental/psychological illness.

| Measure | Estimate |
| --- | --- |
| Weighted prevalence of any reported disability | 7.30% |
| Weighted prevalence of reported mental/psychological illness | 0.71% |
| Observed weighted prevalence of both | 0.5032% |
| Expected prevalence under independence | 0.05185% |
| Observed – expected | 0.45134 percentage points |
| Observed/expected ratio | 9.70 |
| 95% CI for observed/expected ratio | 7.60–11.81 |
| Design-based Pearson test | $F(1,170)=239.54$ |
| p value | $<0.001$ |
| Adjusted association of disability with | OR 42.99 |
| mental/psychological illness |  |
| 95% CI | 18.45–100.18 |
| p value | <0.001 |
| Sensitivity adjusted association | OR 29.68 |
| 95% CI | 12.83–68.66 |
| p value | <0.001 |

**Note:** Expected prevalence under independence was calculated as the product of the two marginal weighted prevalences. The primary association model used reported mental/psychological illness as the outcome and adjusted for any reported disability, age group and sex. The sensitivity model additionally adjusted for marital status and household size. These are cross-sectional associations and do not establish direction or causality.

In the parsimonious survey-weighted logistic model adjusted for age group and sex, adults with reported disability had substantially higher odds of reported mental/psychological illness (OR 42.99, 95% CI 18.45–100.18, ***p***<0.001). After additional adjustment for marital status and household size, the association remained statistically detectable (OR 29.68, 95% CI 12.83–68.66, ***p***<0.001; Table 5; Table S15 in S1 Table).

The four-category co-occurrence distribution varied across education, residence and region. Across education categories, the proportions classified as having both conditions ranged from 0.09% among adults with secondary education to 1.03% among those with primary education; the overall education association was statistically detectable (*p*=0.0022). By residence, the prevalence of the “both” category was 0.58% among urban adults, 0.58% among rural adults and 0.18% among nomadic adults (*p*<0.001). Regional prevalence of the joint category ranged from 0.25% in Sanaag to 0.65% in Togdheer (*p*=0.0031). The complete four-category distributions are provided in Table S16 in S1 Table.

The wealth-stratified co-occurrence prevalence was also non-uniform, ranging from approximately 0.30% in the lowest wealth quintile to 0.91% in the middle quintile (Table S1 in S1 Table). The corresponding concentration index for the joint outcome was -0.0432 (SE 0.0901, ***p***=0.6323), with no statistically detectable marginal socioeconomic concentration (Table S7 in S1 Table).

Because only 68 co-occurrence events were observed, several covariate cells were sparse, particularly among education categories; this limits precision and requires cautious interpretation of detailed regression estimates (Table S11 in S1 Table; Table S6 in S1 Table).

### Domain-specific patterns by sex and age

Overall disability prevalence was 6.93% among men and 7.61% among women, and sex was not independently associated with any reported disability in the primary multivariable model. At the domain level, hearing was the only disability domain with a statistically detectable sex difference, occurring in 1.03% of men and 1.95% of women (*p*=0.0129). The remaining domains did not show statistically detectable sex differences (Table S12 in S1 Table).

Age variation was more pronounced across disability domains. Sight disability increased from 1.73% among adults aged 18-29 years to 14.72% among adults aged ≥60 years. Hearing increased from 0.81% to 6.38%, and mobility from 0.86% to 4.90% across the same age groups. Survey-adjusted age differences were statistically detectable for sight, hearing, learning, mobility, self-care and the mental disability domain, whereas the age difference for speech did not reach conventional statistical significance. Complete domain-specific age estimates are presented in Table S13 in S1 Table.

### Reported disability origin, age at onset and support

Among 799 adults with reported disability and valid origin responses, the most frequently reported origins were aging (30.93%) and other disease (23.81%), followed by contagious disease (10.71%), congenital conditions (10.02%) and injury or accident (9.96%). These were respondent-reported contextual attributions rather than causal classifications (Table S17 in S1 Table).

Among the same 799 adults with valid age-at-onset information, 15.59% reported onset before age 5 years, 5.95% at ages 5–9 years, 11.06% at ages 10–19 years, 11.07% at ages 20–29 years, 6.63% at ages 30–39 years, 12.71% at ages 40–49 years, 10.33% at ages 50–59 years, 13.16% at ages 60-69 years, and 13.50% at age ≥70 years (Table S18 in S1 Table).

Support information was available for 921 of the 947 adults with reported disability. Medical support was reported by 79.95%, no support by 29.86%, financial support by 2.56%, nutritional support by 0.96%, and welfare support by 0.56%. Because these were multiple-response indicators, the percentages were not mutually exclusive (Table S19 in S1 Table).

### Missing data, data-quality and sensitivity analyses

Education was missing for 180 adults (1.23%). Weighted disability prevalence was 7.29% among adults with complete education information and 8.56% among adults with missing education. The association between education missingness and disability was not statistically detectable (OR 1.19, 95% CI 0.49–2.88, *p*=0.698). This analysis does not establish that the education data were missing completely at random (Table S9 in S1 Table).

A source-data inconsistency was identified in 227 adults for whom “None” was recorded alongside at least one positive disability-domain indicator. Using the positive labelled domain indicators as authoritative, the primary prevalence was 7.30%; excluding the 227 conflicting records reduced weighted disability prevalence to 5.13% (95% CI 4.58–5.73). The fully adjusted wealth association remained statistically non-detectable after exclusion (F(4,167)=0.86, *p*=0.491), indicating that the sensitivity analysis did not materially alter the conclusion regarding wealth (Table S20 in S1 Table).

Additional covariate-specification analyses yielded consistent results. Adding marital status and household size to the primary disability model gave a global wealth *p*=0.687, while additionally including relationship to household head gave *p*=0.494 (Table S21 in S1 Table).

## Discussion

### Principal findings

This study provides an adult-focused analysis of reported disability and reported mental/psychological illness using the 2020 Somaliland Health and Demographic Survey (SLHDS). Among 14,579 adults, 7.30% had at least one reported disability domain. Sight, hearing and mobility were the most frequently reported domains, while the separate mental disability domain was reported by 1.29% of adults. Reported disability showed a marked age gradient, with substantially higher prevalence among older adults, and substantial variation across residence and region. The crude association between household wealth and reported disability was attenuated after adjustment for residence and region and was not statistically detectable in the primary multivariable model. Nevertheless, the concentration analysis identified a modest positive marginal concentration of reported disability toward higher wealth ranks. Separately reported mental/psychological illness was uncommon in the survey (0.71%), but its co-occurrence with reported disability was substantially greater than expected under statistical independence.

These findings address the specific gap identified in the study protocol: the SLHDS 2020 report provided all-age descriptive information on disability but did not provide this adult-specific survey-weighted analysis of disability, socioeconomic inequality, or co-occurrence with the separately recorded mental/psychological illness construct (10). The findings should therefore be interpreted as estimates for the 2020 survey period, rather than as current prevalence estimates or temporal trends.

### Magnitude and composition of reported disability

The estimated prevalence of any reported disability among adults was 7.30%. This estimate should not be directly equated with the approximately 16% global estimate reported by WHO because prevalence depends substantially on population age structure, measurement instruments and operational definitions (1,2). Similarly, the present estimate is not a reproduction of the approximately 5% disability estimate reported for the SLHDS population as a whole, because the present analysis uses an explicitly defined adult denominator and applies the survey design to that population (10).

The composition of disability provides additional information beyond the overall prevalence. Sight was the most frequently reported domain, followed by hearing and mobility. This broad predominance of sensory and mobility-related domains is compatible with evidence from sub-

Saharan African household surveys, although direct numerical comparisons should be made cautiously because disability instruments, age structures and operational definitions differ between surveys (3). The seven disability domains were not mutually exclusive, so the domain-specific estimates describe overlapping dimensions of reported disability rather than separate population groups.

The distinction between the mental disability domain and reported mental/psychological illness is particularly important when interpreting these findings. The mental disability domain belongs to the SLHDS disability-domain item set, whereas reported mental/psychological illness was recorded separately in the chronic-disease module (10). The two measures therefore should not be interpreted as interchangeable indicators of the same condition. The difference between their prevalences in this analysis also illustrates why combining them into a single mental-health outcome would obscure the structure of the original survey measures.

### Age and sex patterning

Age was the clearest demographic pattern in the analysis. Reported disability increased from 4.15% among adults aged 18-29 years to 23.96% among adults aged ≥60 years. In the primary adjusted model, adults aged 45–59 years had higher odds of reported disability than those aged 18-29 years, while adults aged ≥60 years had substantially higher odds. The ordered age analysis and the continuous-age sensitivity analysis produced the same overall pattern.

This age gradient is consistent with the broader evidence that disability generally increases with age (1,3). The domain-specific analyses showed that the age pattern was especially pronounced for sight, hearing, mobility and self-care. The results therefore suggest that the overall age gradient was not driven by a single disability domain but reflected variation across several domains.

The cross-sectional design does not establish the mechanism underlying this association. The appropriate interpretation is therefore an observed age-associated distribution rather than an estimate of an age effect.

Sex differences were comparatively limited. Overall reported disability was somewhat higher among females than males, but sex was not independently associated with the primary disability outcome after adjustment. At the domain level, hearing was the only domain for which the sex comparison reached conventional statistical significance. This suggests that sex-related patterning may differ across disability domains rather than producing a uniform difference in overall reported disability.

### Socioeconomic patterning and geographic context

The wealth findings require interpretation of two complementary analyses. In crude analysis, reported disability differed across wealth quintiles, but the pattern was non-monotonic: prevalence was lowest in the lowest quintile and higher in several middle quintiles before declining again in the highest quintile. When demographic and geographic variables were incorporated, the wealth association was progressively attenuated and was not statistically detectable in the primary model containing residence and region.

This pattern indicates that the crude wealth distribution and the conditional regression association answer different questions. Household wealth, residence and region were themselves strongly patterned in the study population, so the adjusted wealth estimates represent comparisons conditional on overlapping geographic and demographic characteristics. The loss of a statistically detectable wealth association after geographic adjustment should therefore not be interpreted as evidence that socioeconomic position is unrelated to disability in general. Rather, within this dataset, the independent association attributed to wealth quintile was sensitive to adjustment for residence and region.

The concentration analysis adds another dimension. The concentration index for reported disability was positive and statistically detectable, indicating relative concentration of the reported outcome toward higher wealth ranks in the marginal population distribution. This does not contradict the null adjusted wealth association. A concentration index describes how the outcome is distributed across the socioeconomic ranking, whereas the multivariable regression estimates conditional associations after adjustment for selected characteristics (4). The two results therefore describe different aspects of socioeconomic patterning.

The positive marginal concentration is also consistent with the observation from multi-country sub-Saharan African evidence that disability does not necessarily show a simple pro-poor gradient across settings (3). At the same time, the present findings should not be interpreted as evidence that higher wealth causes disability or that disability is intrinsically concentrated among wealthier households. Differences in population composition, residence, geography, recognition, reporting and access to services may contribute to the observed distribution.

The self-care domain showed a statistically detectable negative concentration index, suggesting relative concentration toward lower wealth ranks. Because self-care disability was uncommon and the domain-specific estimate was comparatively imprecise, this finding should be treated as a domain-specific distributional observation rather than evidence of a general socioeconomic mechanism.

Geographic heterogeneity persisted after adjustment. Region remained jointly associated with reported disability, with higher adjusted odds in Marodijeh, Sahil, Togdheer and Sool relative to Awdal. Residence also remained jointly associated, with the adjusted difference driven particularly by the nomadic category. These findings demonstrate that national prevalence can conceal meaningful spatial and livelihood-related variation.

The regional and residence findings should not be interpreted as causal effects of belonging to a particular region or residence category. Region and residence represent broad social, geographic and livelihood contexts that may capture differences in population composition, environment, mobility, service access, physical infrastructure, reporting and recognition. The present analysis establishes the existence of these associations but cannot identify their underlying mechanisms.

### Reported mental/psychological illness and disability co-occurrence

Reported mental/psychological illness was uncommon in the survey, with a weighted prevalence of 0.71%. This estimate should be understood in the context of the survey measurement: the outcome was a reported chronic-disease item and was not a standardized psychiatric diagnostic assessment. Its prevalence may therefore reflect, in addition to underlying illness, recognition, access to care, contact with health services and recording of the condition.

Despite its low overall prevalence, the joint occurrence of reported mental/psychological illness and reported disability was notable. The observed prevalence of both conditions was 0.503%, compared with an expected prevalence of approximately 0.052% under statistical independence. The observed-to-expected ratio was 9.70, and the design-based test provided statistical evidence against independence.

The adjusted association analysis was consistent with this result. After adjustment for age group and sex, adults with reported disability had substantially higher odds of reported mental/psychological illness; the association remained statistically detectable after additional adjustment for marital status and household size. These results are consistent in direction with international evidence showing an association between mental and physical conditions and disability (5). However, the present findings are not directly equivalent to those studies because the SLHDS measures were survey-reported constructs obtained from different modules rather than standardized diagnostic assessments.

The large odds ratio should therefore be interpreted specifically as a strong cross-sectional association between two reported survey constructs. It does not establish whether mental/psychological illness precedes disability, whether disability contributes to mental/psychological illness, or whether both are influenced by shared underlying factors. Recognition and access to diagnosis may also influence the observed relationship.

The socioeconomic analysis of co-occurrence should similarly be interpreted cautiously. The four-category distribution varied across education, residence, region and wealth, but the mental/psychological illness-only and both-condition categories contained relatively few observations. The 68 observed co-occurrence events resulted in wide uncertainty around several detailed estimates. Consequently, statistically non-detectable subgroup differences should not be interpreted as proof that no socioeconomic differences exist.

### Contextual profile of disability

The contextual analyses provide information that is not captured by prevalence and regression estimates alone. Among adults with valid origin information, aging and other disease were the most frequently reported origins, followed by contagious disease, congenital conditions and injury or accident. These categories represent respondents’ reported attributions and should not be interpreted as population-level causal fractions.

The reported age-at-onset distribution also demonstrated that disability was not confined to later adulthood. Some respondents reported onset during childhood or adolescence, while others reported onset during middle or older adulthood. This complements the strong cross-sectional age gradient in current disability prevalence: the higher prevalence among older adults does not imply that all disability begins late in life.

Medical support was reported frequently among adults with available support information, but nearly three in ten were recorded as receiving no support. Because the support variables were multiple-response indicators, and because the study did not measure adequacy, continuity, quality or effectiveness of support, these estimates describe recorded support patterns rather than directly measuring unmet need or service effectiveness.

### Data-quality findings and implications for interpretation

An important finding of the analysis was the internal inconsistency involving 227 adults for whom “None” was recorded together with at least one positive disability-domain indicator. The primary analysis used the labelled positive domain indicators as authoritative, while exclusion of the conflicting observations reduced the estimated disability prevalence from 7.30% to 5.13%.

The persistence of the non-detectable wealth association in the sensitivity analysis indicates that the principal inference concerning wealth was not materially changed by this coding issue.

Nevertheless, the magnitude of the prevalence change demonstrates that the disability estimate is partly dependent on the adopted rule for resolving this source-data inconsistency. This should be reported transparently rather than hidden in the supplementary analysis. It also emphasizes the importance of distinguishing the survey’s recorded disability indicators from clinical diagnostic measures.

The missing-education analysis showed that education was missing for only 1.23% of adults, and there was no statistical evidence that missing education was associated with reported disability. However, this does not establish that the data were missing completely at random. The primary models therefore appropriately retained complete-case analyses rather than imposing an unsupported imputation mechanism.

### Strengths and limitations

This analysis has several strengths. First, it uses the adult household-member population as an explicit denominator rather than relying on an all-age survey estimate. Second, all inferential analyses incorporate the complex survey structure through weights, stratification and clustering. Third, the analysis preserves all seven recorded disability domains and distinguishes the mental disability domain from the separately recorded mental/psychological illness construct. Fourth, the study combines prevalence estimation, multivariable regression, concentration-based inequality analysis and formal examination of co-occurrence rather than relying on a single measure of disability burden. Finally, the reopened analyses provide domain-specific age and sex patterns and contextual information on origin, onset and support that complement the principal outcomes.

Several limitations must also be considered. The cross-sectional design prevents assessment of temporality or causal direction. Disability and reported mental/psychological illness were survey measures and were not independently clinically validated. The audited disability data did not provide a severity scale that justified applying an external disability threshold, so Washington Group classification was not imposed. Reported mental/psychological illness was based on a survey chronic-disease item and should not be interpreted as equivalent to a standardized psychiatric diagnosis. The small number of mental/psychological illness and co-occurrence events limited precision in detailed regression analyses. Education was missing for 180 adults, leading to complete-case analyses in models that included education. The 227-record “None” plus positive-domain inconsistency introduced sensitivity into the absolute prevalence estimate. Contextual origin and onset variables were available only for respondents with valid responses, and support measures were multiple-response indicators. Finally, because the SLHDS was conducted in 2020, these findings describe the survey period and should not automatically be interpreted as estimates of disability prevalence in 2026.

### Implications for research and population-health planning

The results establish an adult population baseline for reported disability in Somaliland and show that a single national prevalence figure does not capture the distribution of disability across age, residence and region. The marked increase in reported disability among older adults identifies ageing as an important dimension for future disability research within Somaliland, while the regional and residence differences indicate the need for further investigation of geographic and livelihood-related determinants.

The co-occurrence findings also identify a subgroup that warrants more focused investigation. Future studies should use validated mental-health measures, clearer diagnostic ascertainment and more detailed information on healthcare access, recognition and support to determine whether the strong cross-sectional association observed here persists when the limitations of the survey measures are reduced. Longitudinal or repeated cross-sectional data would also be needed to examine changes over time and to distinguish changing burden from changes in recognition or reporting.

The present findings can therefore serve as a baseline for subsequent work using newer Somaliland data, while preserving a clear distinction between what the SLHDS can establish— population distributions and associations—and what requires clinical assessment or longitudinal research.

## Conclusion

In this national adult analysis of SLHDS 2020, 7.30% of adults had at least one reported disability, with sight, hearing and mobility the most frequently reported domains. Reported disability showed a marked age gradient and substantial geographic heterogeneity. The crude association between household wealth and reported disability was attenuated after adjustment for residence and region and was not statistically detectable in the primary multivariable model, while concentration analysis showed a modest positive marginal concentration across the wealth ranking. Separately reported mental/psychological illness was uncommon, but its co-occurrence with reported disability substantially exceeded that expected under statistical independence and remained strongly associated after adjustment for demographic factors.

Taken together, the findings support an interpretation of adult disability in Somaliland centered on age, geographic context and careful separation of disability and mental/psychological illness constructs, while recognizing the limitations of cross-sectional, survey-reported measures (1–5,10).

## Supporting information

S1 Checklist STROBE

S1 Protocol Study and Statistical Analysis SLHDS2020

## Data Availability

The underlying Somaliland Health and Demographic Survey 2020 (SLHDS 2020) microdata are third-party data held by the Central Statistics Department (CSD) of the Ministry of Planning and National Development, Somaliland, and cannot be publicly shared by the authors because access is subject to the data provider's permission and conditions. Permission to access and analyse the anonymized SLHDS 2020 dataset for this study was obtained from the CSD. Researchers seeking access should contact the CSD through its official data-access process. The Stata analysis code used in this study is publicly available and archived in Zenodo: https://doi.org/10.5281/zenodo.22906585.

https://www.somalilandcsd.org/somaliland-health-and-demography-survey-slhd2020/

## Acknowledgments

The authors gratefully acknowledge the Somaliland Ministry of Health Development and the Somaliland Health and Demographic Survey (SLHDS) program for granting access to the 2020 dataset used in this study. The authors deeply appreciate the individuals and households who participated in the survey, whose contributions made this research possible. The authors also thank the data collection teams and the institutions involved in the design, implementation, quality assurance, and management of SLHDS 2020.

## Supporting information

S1 Table. Supplementary Tables. Supplementary analyses supporting the manuscript, including survey-weighted prevalence estimates, regression analyses, concentration indices, missing-data assessment, sensitivity analyses, age- and sex-specific disability estimates, co-occurrence analyses, and contextual disability characteristics.

S2 Checklist. STROBE checklist for this cross-sectional study.

S1 Protocol. Study and statistical analysis protocol for the secondary analysis of the Somaliland Health and Demographic Survey 2020.

