## Supplementary material for "Adult disability and mental/psychological illness in Somaliland: socioeconomic inequalities and co-occurrence in the 2020 Somaliland Health and Demographic Survey": S1 Checklist STROBE

**STROBE checklist for the cross-sectional study**

**Checklist scope.** This completed checklist maps the manuscript to the STROBE cross-sectional reporting items. Reporting elements are concise paraphrases of the official STROBE checklist. Location references use section/subsection names and paragraph numbers. Paragraph numbering restarts at 1 within each named subsection and excludes headings, table captions, figure captions, and notes. Tables and figures are cited by their manuscript labels. This checklist is a reporting map, not a measure of methodological quality.

| **STROBE section** | **Item** | **Reporting element** | **Location in latest manuscript** | **Status** |
| --- | --- | --- | --- | --- |
| Title and abstract | 1a | Study design stated in title or abstract | Abstract, paragraph 2: the study is explicitly described as a cross-sectional secondary analysis. | **Reported** |
| Title and abstract | 1b | Informative, balanced summary of what was done and found | Abstract, paragraphs 1–4: objective, design, population, measures, analyses, principal findings, and limitations are summarized. | **Reported** |
| Introduction | 2 | Scientific background and rationale | Introduction, paragraphs 1–10: global and regional disability context, socioeconomic patterning, mental/psychological illness, Somaliland evidence gap, and rationale. | **Reported** |
| Introduction | 3 | Specific objectives and prespecified hypotheses | Introduction, paragraph 10: study aims are explicitly stated, including the prespecified expectation regarding co-occurrence beyond statistical independence. | **Reported** |
| Methods | 4 | Key design elements stated early | Methods — Study design and data source, paragraph 1: cross-sectional secondary analysis and source survey design are stated at the beginning of Methods. | **Reported** |
| Methods | 5 | Setting, location, and relevant dates | Methods — Study design and data source, paragraph 1: Somaliland, six regions, three residence domains, and 2018–2019 data collection are described. | **Reported** |
| Methods | 6 | Eligibility criteria and participant selection | Methods — Study population and analytic sample, paragraphs 1–3: source records, adult eligibility, usual-resident criterion, disability-module status, chronic-disease gate, exclusions, and final analytic sample are specified. | **Reported** |
| Methods | 7 | Outcomes, exposures, predictors, and potential confounders | Methods — Definition of reported disability, paragraphs 1–4; Definition of reported mental/psychological illness, paragraphs 1–2; Disability–mental/psychological illness co-occurrence, paragraphs 1–6; Socioeconomic and demographic variables, paragraphs 1–3. | **Reported** |
| Methods | 8 | Sources of data and methods of measurement | Methods — Study design and data source, paragraph 1; Definition of reported disability, paragraphs 1–2; Definition of reported mental/psychological illness, paragraphs 1–2; contextual disability variables, paragraphs 1–3. | **Reported** |
| Methods | 9 | Potential sources of bias | Methods — Definition of reported disability, paragraph 4; Contextual and sensitivity analyses, paragraphs 2–3; Missing data, paragraphs 1–2. Measurement, sparse-event, missing-data, and source-data limitations are also discussed in Discussion — Strengths and limitations, paragraphs 1–2. | **Reported** |
| Methods | 10 | How study size was determined | Methods — Study population and analytic sample, paragraph 2: all eligible records available in the existing survey were included and no a priori sample-size or power calculation was performed. | **Reported** |
| Methods | 11 | Handling of quantitative variables | Methods — Socioeconomic and demographic variables, paragraphs 1–3; Contextual disability variables, paragraphs 1–3; Statistical analysis — Socioeconomic distribution and multivariable analysis of disability, paragraph 2. Age boundaries, wealth ranking, continuous-age sensitivity analysis, and contextual groupings are specified. | **Reported** |
| Methods | 12a | Statistical methods, including confounding control | Methods — Statistical analysis, paragraph 1; Descriptive analysis, paragraphs 1–2; Socioeconomic distribution and multivariable analysis of disability, paragraphs 1–3; Socioeconomic inequality analysis, paragraphs 1–2; Mental/psychological illness analysis, paragraphs 1–2; Co-occurrence and independence analysis, paragraphs 1–2; Contextual and sensitivity analyses, paragraphs 1–3; Missing data, paragraphs 1–2. | **Reported** |
| Methods | 12b | Subgroup and interaction methods | Methods — Statistical analysis — Descriptive analysis, paragraph 2: disability domains are examined by sex and age group and the ordered age gradient is assessed. No interaction analyses were performed. | **Reported** |
| Methods | 12c | Missing-data handling | Methods — Missing data, paragraphs 1–2: education missingness is quantified, complete-case analyses are used for models requiring education, and no imputation is performed. | **Reported** |
| Methods | 12d | Analytical methods accounting for sampling strategy | Methods — Survey design, weights, and complex-sample estimation, paragraphs 1–2; Statistical analysis, paragraph 1: sampling weights, 18 strata, 188 stratum-specific PSUs, clustering, and linearized variance are specified. | **Reported** |
| Methods | 12e | Sensitivity analyses | Methods — Contextual and sensitivity analyses, paragraphs 1–3: conflict-record exclusion, alternative age specification, additional covariate specifications, and limitations of unreconstructable source-level analyses are documented. | **Reported** |
| Results | 13a–c | Participant numbers at each stage and flow | Results — Analytic population, paragraph 1; Fig 1: source count, adult count, exclusions, final N=14,579, and analytical flow are reported. | **Reported** |
| Results | 14a–b | Participant characteristics and missingness | Results — Analytic population, paragraph 2; Table 1; Results — Missing data, data-quality and sensitivity analyses, paragraph 1. | **Reported** |
| Results | 15 | Outcome events and summary measures | Results — Prevalence and composition of reported disability, paragraphs 1–2; Co-occurrence of reported disability and reported mental/psychological illness, paragraphs 1–2; Tables 2 and 5. | **Reported** |
| Results | 16a | Unadjusted/adjusted estimates with precision | Results — Socioeconomic and geographic distribution of reported disability, paragraphs 1–4; Multivariable association between household wealth and reported disability, paragraphs 1–2; Primary multivariable model, paragraph 1; Co-occurrence, paragraphs 2–4; Tables 3–5. | **Reported** |
| Results | 16b | Category boundaries for categorized quantitative variables | Methods — Socioeconomic and demographic variables, paragraph 3; Results — Age pattern, paragraphs 1–2. The age groups 18–29, 30–44, 45–59, and ≥60 years are explicitly defined and used. | **Reported** |
| Results | 16c | Translation of relative measures to absolute risk when relevant | Not applicable: the study is a cross-sectional prevalence/association analysis and does not estimate longitudinal risk. | **N/A** |
| Results | 17 | Other analyses, subgroup, and sensitivity analyses | Results — Age pattern, paragraph 2; Concentration-based socioeconomic inequality, paragraphs 1–2; Reported mental/psychological illness, paragraphs 1–3; Co-occurrence, paragraphs 2–4; Domain-specific patterns by sex and age, paragraphs 1–2; Reported disability origin, age at onset and support, paragraphs 1–3; Missing data, data-quality and sensitivity analyses, paragraphs 1–3. | **Reported** |
| Discussion | 18 | Key results linked to objectives | Discussion — Principal findings, paragraphs 1–2. | **Reported** |
| Discussion | 19 | Limitations, bias, and imprecision | Discussion — Strengths and limitations, paragraphs 1–2; Data-quality findings and implications for interpretation, paragraphs 1–3. | **Reported** |
| Discussion | 20 | Cautious interpretation in light of limitations and evidence | Discussion — Magnitude and composition of reported disability, paragraphs 1–3; Age and sex patterning, paragraphs 1–4; Socioeconomic patterning and geographic context, paragraphs 1–7; Reported mental/psychological illness and disability co-occurrence, paragraphs 1–5. | **Reported** |
| Discussion | 21 | Generalisability/external validity | Methods — Study design and data source, paragraph 1; Discussion — Strengths and limitations, paragraph 2; Implications for research and population-health planning, paragraphs 1–3. The scope is restricted to the 2020 survey period and recorded constructs. | **Reported** |
| Other information | 22 | Funding source and role of funder | PLOS submission system — Financial Disclosure/Funding section. The manuscript does not include a funding section. | **Reported in submission system — not in manuscript** |

**Source note:** STROBE cross-sectional reporting items are paraphrased for this checklist. The location references were reconstructed directly from the manuscript structure.
