## Supplementary material for "Adult disability and mental/psychological illness in Somaliland: socioeconomic inequalities and co-occurrence in the 2020 Somaliland Health and Demographic Survey": S1 Protocol Study and Statistical Analysis SLHDS2020

**Study and statistical analysis protocol for the secondary analysis of the Somaliland Health and Demographic Survey 2020**

**Adult disability and mental/psychological illness in Somaliland: socioeconomic inequalities and co-occurrence in the 2020 Somaliland Health and Demographic Survey**

### 1. Study design

This study was a cross-sectional secondary analysis of the 2020 Somaliland Health and Demographic Survey (SLHDS 2020), a nationally representative household survey conducted in Somaliland. The source survey used a stratified multistage probability cluster design covering six regions and three residence domains: urban, rural, and nomadic. Disability and chronic-disease information was collected through the household questionnaire for household members rather than through clinical examination.

The SLHDS main survey data collection in urban and rural areas was conducted from August 2018 to December 2019, with nomadic data collection conducted as part of the corresponding main-survey fieldwork. The analysis was restricted to adults aged 18 years or older and interpreted all estimates as distributions or associations rather than causal effects because the source survey was cross-sectional.

### 2. Study population and eligibility

The source person-level file contained 35,381 household-member records. The analytic population was restricted to adults aged ≥18 years who:

- were usual residents (HV102=1);
- had the disability module administered; and
- had a defined reported mental/psychological illness measure based on an answered chronic-disease screening gate.

Among 15,041 adults with usable age information, 342 non-usual residents were excluded, six adults were excluded because the disability module was not administered, and 114 were excluded because the chronic-disease gate was coded “Don't know.” The final analytic sample was therefore 14,579 adults. Because this was a secondary analysis of an existing population-based survey, the analytic sample comprised all eligible records available in the source dataset; no a priori sample-size or power calculation was performed.

The analysis used the de jure/usual-resident population rather than the de facto population. A de facto adult count of 14,317 was retained solely as a data-preparation reference and was not used as the denominator for the primary or secondary analyses.

### 3. Survey design, weights, and complex-sample estimation

All inferential analyses incorporated the SLHDS sampling design through sampling weights, stratification, and clustering. The final analytic dataset contained 18 sampling strata, corresponding to the six regions crossed by the three residence domains, and 188 unique stratum-specific primary sampling-unit (PSU) units.

The underlying cluster identifier (HV001) was not unique across strata, so the analytical PSU was represented at the stratum-specific level. The survey declaration used the final analysis weight, PSU, and stratum variables, with single-unit(centered) specified for strata containing a single sampled PSU. The same survey declaration was retained throughout the analysis.

### 4. Primary outcome: reported disability

Reported disability was the primary outcome and was constructed from seven SLHDS disability-domain indicators:

- sight
- hearing
- speech
- learning
- mobility
- self-care
- mental

Any positive response in at least one of the seven domains classified an adult as having any reported disability. The seven domains were retained as separate secondary outcomes and were allowed to overlap. No external severity threshold or Washington Group classification was imposed because the audited SLHDS materials did not provide a graded severity scale or establish use of the Washington Group Short Set.

Because the concatenated HV408 multi-response field was width-limited, the labelled domain-specific indicators were treated as authoritative. An internal inconsistency involving 227 adults with both “None” and a positive disability domain was retained for quality control and evaluated in sensitivity analysis.

### 5. Separate mental/psychological illness outcome

Reported mental/psychological illness was treated as a distinct secondary outcome and was not derived from the mental disability domain. It was constructed from chronic-disease indicator HV402Q, conditional on the chronic-disease screening gate HV401. A positive outcome required HV401=1 and HV402Q=1; a negative value was assigned when the chronic-disease gate was positive but HV402Q was not selected, or when HV401=2. Records with an unresolved “Don't know” chronic-disease response were excluded from the final analytic population.

The physician-information variable HV403 was not used to define the outcome. The manuscript therefore refers to this construct as “reported mental/psychological illness” rather than physician-diagnosed mental illness.

### 6. Co-occurrence definition

Reported disability and reported mental/psychological illness were cross-classified into four mutually exclusive groups:

1. Neither: no reported disability and no reported mental/psychological illness.
2. Disability only: reported disability without reported mental/psychological illness.
3. Mental/psychological illness only: reported mental/psychological illness without reported disability.
4. Both conditions: reported disability and reported mental/psychological illness.

The two constructs were kept separate and were not combined into a clinical diagnosis.

### 7. Exposures and covariates

The primary socioeconomic exposure was household wealth index quintile, an asset-based relative measure. The continuous wealth-index factor score was used as the ranking variable for concentration-based inequality analysis.

Secondary socioeconomic/contextual variables were:

- education
- residence
- region

Demographic and social covariates included:

- age
- sex
- marital status
- household size
- relationship to household head

Age was analysed both categorically and continuously. The primary disability model used four age groups: 18–29, 30–44, 45–59, and ≥60 years. Education was classified as no education, primary, secondary, or higher education; the non-formal “Koranic” category was retained as missing rather than incorporated into formal schooling categories. Residence was categorized as urban, rural, or nomadic, and region as Awdal, Marodijeh, Sahil, Togdheer, Sool, or Sanaag.

### 8. Contextual disability variables

Among adults with reported disability, the analysis additionally described reported origin of disability, age at onset, and care/support received. Origin categories followed the source coding and included congenital, contagious disease, child-birth conditions, other disease, abuse, aging, injury/accident, witchcraft, other, and don't know.

Age at onset was grouped as <5, 5–9, 10–19, 20–29, 30–39, 40–49, 50–59, 60–69, and ≥70 years. Care/support variables covered medical, welfare, financial, nutritional, and no-support responses. Support variables were treated as multiple-response indicators, so percentages were not expected to sum to 100%.

### 9. Statistical analysis

All analyses were conducted using Stata/MP version 17.0 (StataCorp, College Station, TX, USA) using the svy framework with linearized variance estimation. Survey-adjusted 95% confidence intervals were reported unless otherwise specified. All hypothesis tests were two-sided, and p<0.05 was considered statistically significant. No formal adjustment for multiple comparisons was applied; domain-specific, sex- and age-stratified, and other secondary analyses were interpreted as exploratory, and their p values were not treated as evidence from independent confirmatory hypotheses.

#### 9.1 Descriptive analysis

Survey-weighted proportions were estimated for any reported disability; each of the seven disability domains; reported mental/psychological illness; and the four co-occurrence categories. Domain-specific disability was examined by sex and age group using design-based tests. The four age groups were also used to evaluate the prespecified age gradient.

#### 9.2 Socioeconomic distribution and regression

Disability prevalence was estimated across wealth quintiles, education, residence, and region. Survey-weighted logistic regression was used to examine associations between reported disability and socioeconomic/demographic variables.

The primary multivariable model simultaneously included household wealth quintile, age group, sex, education, residence, and region. Alternative specifications examined the wealth association under different adjustment sets. Adjusted predicted probabilities were also estimated by wealth quintile. Sensitivity models additionally included marital status, household size, and relationship to household head.

#### 9.3 Concentration-based socioeconomic inequality

Socioeconomic inequality was assessed using a concentration index, ranking individuals by the continuous household wealth-index factor score from lower to higher wealth. Because disability is a bounded binary outcome, the Erreygers normalized concentration index was also estimated. Standard errors and statistical inference incorporated sampling weights, stratification, and clustering. The concentration analysis was interpreted as a marginal distributional measure and was not treated as equivalent to the adjusted regression association.

#### 9.4 Mental/psychological illness analysis

Weighted prevalence of reported mental/psychological illness was estimated overall and across age, sex, education, residence, region, and wealth. Survey-weighted logistic regression examined its association with socioeconomic and demographic variables. Because there were only 99 positive events, interpretation emphasized the resulting imprecision and wide confidence intervals.

#### 9.5 Co-occurrence and independence analysis

The four-category co-occurrence outcome was examined across wealth, education, residence, and region using survey-weighted descriptive comparisons and design-based tests. A survey-weighted multinomial logistic model was used for the categorical wealth analysis.

To assess departure from independence, the expected joint prevalence was calculated as P(disability) × P(mental/psychological illness). The observed-to-expected ratio was calculated as P(both) ÷ [P(disability) × P(mental/psychological illness)]. The ratio and its 95% confidence interval were estimated using Stata's nonlinear combination (nlcom) procedure, propagating uncertainty from the estimated marginal and joint prevalences.

A survey-weighted logistic model then examined the association between disability and reported mental/psychological illness, adjusted for age group and sex. A sensitivity model additionally adjusted for marital status and household size. These models were interpreted as cross-sectional associations, not causal effects.

#### 9.6 Sensitivity and data-quality analyses

The analysis included:

- exclusion of the 227 records with “None” plus a positive disability-domain response;
- alternative wealth-adjustment specifications;
- continuous-age sensitivity analysis;
- additional adjustment for marital status and household size;
- additional adjustment for relationship to household head; and
- assessment of education missingness.

No statistical imputation was performed. Education was missing for 180 adults (1.23%), and analyses requiring education used complete observations. The frozen analytic dataset did not retain sufficient information to reconstruct certain earlier proposed sensitivity analyses; those analyses were therefore not represented as completed analyses.

### 10. Missing data

The core analysis variables—including reported disability, reported mental/psychological illness, co-occurrence, survey weight, PSU, and stratum—were complete in the final analytic dataset. Missingness was concentrated in education and contextual disability variables. The primary multivariable analysis therefore used complete observations for the variables required by each model, and no missing-value imputation was introduced.

### 11. Ethical and data-access considerations

This study was a secondary analysis of anonymized data from the 2020 Somaliland Health and Demographic Survey (SLHDS). Ethical approval for the original SLHDS was obtained by the Central Statistics Department (CSD), Ministry of Planning and National Development, in collaboration with the Ministry of Health Development (MoHD), Republic of Somaliland, before data collection. Permission to access and analyse the anonymized SLHDS dataset for this study was obtained from the Central Statistics Department. Because the present study involved secondary analysis of fully de-identified data, no additional ethical approval or informed consent from participants was required.

### 12. Protocol-to-manuscript alignment note

This supporting protocol is aligned to the latest manuscript version and reproduces the study design, analytic population, variable definitions, survey-design specification, primary and secondary analyses, co-occurrence procedure, and completed sensitivity/data-quality analyses reported in that manuscript. It does not represent analyses that were not completed in the frozen analytic dataset.
