## Supplementary material for "Adult disability and mental/psychological illness in Somaliland: socioeconomic inequalities and co-occurrence in the 2020 Somaliland Health and Demographic Survey": S1 Table Supplementary Tables

Note: These supplementary tables are aligned to the analytic dataset (N=14,579). Survey-weighted inferential analyses account for the final survey weight, stratification and clustering. The analytic design contains 18 sampling strata and 188 stratum-specific PSU units. Reported disability is based on the seven SLHDS disability-domain indicators. The mental/psychological illness measure is the separately recorded chronic-disease item and is kept distinct from the mental disability domain. Where a model includes education, the complete-case sample is N=14,399.

Model-diagnostic note: A collinearity diagnostic was performed for the primary adjusted disability specification; the mean variance inflation factor (VIF) was 1.75 and the maximum VIF was 3.71. No coefficient-level outlier removal or winsorization is represented in the supplied analysis code.

Note: Regression-reporting revision. Regression tables report the coefficient on the log-odds scale (β), its standard error (SE), odds ratio (OR), 95% confidence interval (CI), and P value. Model-level overall Wald tests and global wealth tests are reported when available. Survey-model goodness-of-fit statistics are included when directly verified in Stata; where an executed output was not retained, the GOF statistic is not invented.

**Table S1. Survey-weighted prevalence of reported disability, reported mental/psychological illness, and co-occurrence by household wealth quintile**

| **Outcome** | **Lowest** | **Second** | **Middle** | **Fourth** | **Highest** |
| --- | --- | --- | --- | --- | --- |
| Any reported disability (%) | 5.10 (3.96–6.24) | 9.10 (7.01–11.20) | 9.16 (7.56–10.75) | 8.55 (6.74–10.36) | 7.33 (5.92–8.73) |
| Reported mental/psychological illness (%) | 0.491 (0.342–0.705) | 1.204 (0.759–1.905) | 1.279 (0.706–2.305) | 0.780 (0.307–1.965) | 0.521 (0.164–1.644) |
| Disability + mental/psychological illness (%) | 0.310 (0.192–0.501) | 0.842 (0.454–1.558) | 0.914 (0.436–1.906) | 0.499 (0.223–1.111) | 0.435 (0.109–1.720) |

Note: Values are survey-weighted prevalence estimates with 95% CIs. These are descriptive/marginal estimates, not adjusted effects. Wealth quintile 1 (Lowest) is the reference socioeconomic group for regression analyses.

**Table S2. Univariable survey-weighted logistic regression for any reported disability**

| **Term** | **β (log odds)** | **SE** | **OR** | **95% CI** | **P** |
| --- | --- | --- | --- | --- | --- |
| Household wealth: Second vs lowest | 0.621 | 0.188 | 1.86 | 1.29–2.70 | 0.001 |
| Household wealth: Middle vs lowest | 0.631 | 0.157 | 1.88 | 1.38–2.55 | <0.001 |
| Household wealth: Fourth vs lowest | 0.554 | 0.169 | 1.74 | 1.25–2.42 | 0.001 |
| Household wealth: Highest vs lowest | 0.385 | 0.157 | 1.47 | 1.08–2.00 | 0.014 |
| Age group: 30–44 vs 18–29 | 0.122 | 0.158 | 1.13 | 0.83–1.54 | 0.435 |
| Age group: 45–59 vs 18–29 | 0.464 | 0.195 | 1.59 | 1.08–2.32 | 0.019 |
| Age group: ≥60 vs 18–29 | 1.984 | 0.145 | 7.27 | 5.48–9.66 | <0.001 |
| Sex: Female vs male | 0.104 | 0.094 | 1.11 | 0.92–1.33 | 0.271 |
| Education: Primary vs none | -0.342 | 0.185 | 0.71 | 0.49–1.01 | 0.059 |
| Education: Secondary vs none | -0.755 | 0.215 | 0.47 | 0.31–0.72 | 0.001 |
| Education: Higher vs none | -0.635 | 0.235 | 0.53 | 0.33–0.83 | 0.007 |
| Residence: Rural vs urban | -0.062 | 0.110 | 0.94 | 0.76–1.17 | 0.587 |
| Residence: Nomadic vs urban | -0.892 | 0.248 | 0.41 | 0.25–0.66 | <0.001 |
| Region: Marodijeh vs Awdal | 0.582 | 0.151 | 1.79 | 1.33–2.40 | <0.001 |
| Region: Sahil vs Awdal | 0.351 | 0.168 | 1.42 | 1.02–1.97 | 0.038 |
| Region: Togdheer vs Awdal | 0.536 | 0.157 | 1.71 | 1.26–2.33 | 0.001 |
| Region: Sool vs Awdal | 0.385 | 0.172 | 1.47 | 1.05–2.06 | 0.024 |
| Region: Sanaag vs Awdal | -0.105 | 0.157 | 0.90 | 0.66–1.22 | 0.502 |
| Marital status: Divorced vs married | 0.489 | 0.185 | 1.63 | 1.13–2.33 | 0.009 |
| Marital status: Abandoned vs married | 1.322 | 0.366 | 3.75 | 1.83–7.69 | <0.001 |
| Marital status: Widowed vs married | 1.867 | 0.116 | 6.47 | 5.15–8.11 | <0.001 |
| Marital status: Never married vs married | 0.432 | 0.128 | 1.54 | 1.20–1.98 | 0.001 |

Note: Each predictor represents a separate univariable survey-weighted logistic regression. Education analysis used N=14,399; the other models used N=14,579. Reference categories are shown in the term labels. β and SE are on the log-odds scale.

**Table S3. Survey-weighted logistic regression models examining household wealth and reported disability under alternative adjustment sets**

**Model 1: age + sex + education**

| Term | β (log odds) | SE | OR | 95% CI | P |
| --- | --- | --- | --- | --- | --- |
| Wealth: Second vs lowest | 0.635 | 0.211 | 1.887 | 1.244–2.864 | 0.003 |
| Wealth: Middle vs lowest | 0.699 | 0.169 | 2.013 | 1.443–2.807 | <0.001 |
| Wealth: Fourth vs lowest | 0.642 | 0.176 | 1.900 | 1.342–2.691 | <0.001 |
| Wealth: Highest vs lowest | 0.626 | 0.170 | 1.870 | 1.336–2.618 | <0.001 |
| Age: 30–44 vs 18–29 | 0.081 | 0.164 | 1.084 | 0.785–1.499 | 0.622 |
| Age: 45–59 vs 18–29 | 0.456 | 0.201 | 1.577 | 1.061–2.344 | 0.024 |
| Age: ≥60 vs 18–29 | 1.945 | 0.156 | 6.997 | 5.147–9.511 | <0.001 |
| Female vs male | 0.031 | 0.098 | 1.032 | 0.851–1.251 | 0.750 |
| Education: Primary vs none | 0.069 | 0.161 | 1.071 | 0.780–1.471 | 0.670 |
| Education: Secondary vs none | -0.355 | 0.232 | 0.701 | 0.443–1.109 | 0.128 |
| Education: Higher vs none | -0.308 | 0.297 | 0.735 | 0.409–1.320 | 0.301 |
| Constant | -3.580 | 0.161 | 0.028 | 0.020–0.038 | <0.001 |

N=14,399; overall model F(11,160)=38.19, p<0.001; global wealth Wald F(4,167)=5.09, p=0.0007; survey-model GOF F(9,162)=3.69, p=0.0003.

**Model 2: + region**

| Term | β (log odds) | SE | OR | 95% CI | P |
| --- | --- | --- | --- | --- | --- |
| Wealth: Second vs lowest | 0.562 | 0.216 | 1.755 | 1.146–2.686 | 0.010 |
| Wealth: Middle vs lowest | 0.622 | 0.173 | 1.863 | 1.324–2.621 | <0.001 |
| Wealth: Fourth vs lowest | 0.532 | 0.180 | 1.703 | 1.194–2.427 | 0.003 |
| Wealth: Highest vs lowest | 0.440 | 0.189 | 1.553 | 1.068–2.257 | 0.021 |
| Age: 30–44 vs 18–29 | 0.078 | 0.165 | 1.081 | 0.781–1.496 | 0.637 |
| Age: 45–59 vs 18–29 | 0.461 | 0.202 | 1.585 | 1.064–2.361 | 0.024 |
| Age: ≥60 vs 18–29 | 1.964 | 0.160 | 7.131 | 5.204–9.770 | <0.001 |
| Female vs male | 0.020 | 0.099 | 1.020 | 0.839–1.240 | 0.841 |
| Education: Primary vs none | 0.060 | 0.160 | 1.062 | 0.774–1.456 | 0.708 |
| Education: Secondary vs none | -0.361 | 0.233 | 0.697 | 0.440–1.105 | 0.124 |
| Education: Higher vs none | -0.311 | 0.300 | 0.733 | 0.405–1.325 | 0.302 |
| Region: Marodijeh vs Awdal | 0.645 | 0.157 | 1.907 | 1.398–2.600 | <0.001 |
| Region: Sahil vs Awdal | 0.470 | 0.168 | 1.600 | 1.149–2.230 | 0.006 |
| Region: Togdheer vs Awdal | 0.600 | 0.162 | 1.821 | 1.322–2.510 | <0.001 |
| Region: Sool vs Awdal | 0.492 | 0.175 | 1.636 | 1.157–2.313 | 0.006 |
| Region: Sanaag vs Awdal | -0.049 | 0.167 | 0.952 | 0.685–1.323 | 0.768 |
| Constant | -3.952 | 0.209 | 0.019 | 0.013–0.029 | <0.001 |

N=14,399; overall model F(16,155)=30.51, p<0.001; global wealth Wald F(4,167)=3.49, p=0.0091; survey-model GOF F(9,162)=1.38, p=0.2011.

**Model 3: + residence instead of region**

| Term | β (log odds) | SE | OR | 95% CI | P |
| --- | --- | --- | --- | --- | --- |
| Wealth: Second vs lowest | 0.161 | 0.263 | 1.175 | 0.699–1.974 | 0.542 |
| Wealth: Middle vs lowest | 0.138 | 0.262 | 1.148 | 0.684–1.927 | 0.599 |
| Wealth: Fourth vs lowest | 0.030 | 0.271 | 1.030 | 0.603–1.760 | 0.913 |
| Wealth: Highest vs lowest | -0.006 | 0.280 | 0.994 | 0.572–1.727 | 0.983 |
| Age: 30–44 vs 18–29 | 0.062 | 0.164 | 1.063 | 0.769–1.470 | 0.708 |
| Age: 45–59 vs 18–29 | 0.430 | 0.201 | 1.537 | 1.034–2.283 | 0.034 |
| Age: ≥60 vs 18–29 | 1.914 | 0.159 | 6.778 | 4.956–9.270 | <0.001 |
| Female vs male | 0.007 | 0.099 | 1.007 | 0.828–1.225 | 0.943 |
| Education: Primary vs none | 0.012 | 0.159 | 1.012 | 0.739–1.386 | 0.939 |
| Education: Secondary vs none | -0.415 | 0.234 | 0.660 | 0.416–1.048 | 0.078 |
| Education: Higher vs none | -0.365 | 0.296 | 0.694 | 0.387–1.245 | 0.219 |
| Residence: Rural vs urban | -0.276 | 0.145 | 0.759 | 0.570–1.010 | 0.059 |
| Residence: Nomadic vs urban | -1.013 | 0.278 | 0.363 | 0.210–0.629 | <0.001 |
| Constant | -2.869 | 0.300 | 0.057 | 0.031–0.103 | <0.001 |

N=14,399; overall model F(13,158)=37.72, p<0.001; global wealth Wald F(4,167)=0.53, p=0.7132; survey-model GOF F(9,162)=1.29, p=0.2455.

**Model 4: + region + residence**

| Term | β (log odds) | SE | OR | 95% CI | P |
| --- | --- | --- | --- | --- | --- |
| Wealth: Second vs lowest | 0.183 | 0.268 | 1.200 | 0.707–2.039 | 0.497 |
| Wealth: Middle vs lowest | 0.162 | 0.282 | 1.175 | 0.673–2.052 | 0.568 |
| Wealth: Fourth vs lowest | 0.049 | 0.296 | 1.050 | 0.586–1.883 | 0.869 |
| Wealth: Highest vs lowest | -0.031 | 0.311 | 0.970 | 0.525–1.790 | 0.922 |
| Age: 30–44 vs 18–29 | 0.066 | 0.165 | 1.068 | 0.772–1.478 | 0.688 |
| Age: 45–59 vs 18–29 | 0.444 | 0.202 | 1.559 | 1.047–2.321 | 0.029 |
| Age: ≥60 vs 18–29 | 1.939 | 0.162 | 6.949 | 5.051–9.561 | <0.001 |
| Female vs male | -0.001 | 0.101 | 0.999 | 0.819–1.219 | 0.994 |
| Education: Primary vs none | 0.017 | 0.159 | 1.017 | 0.743–1.392 | 0.916 |
| Education: Secondary vs none | -0.405 | 0.234 | 0.667 | 0.420–1.059 | 0.085 |
| Education: Higher vs none | -0.350 | 0.300 | 0.705 | 0.390–1.274 | 0.245 |
| Residence: Rural vs urban | -0.218 | 0.146 | 0.804 | 0.602–1.074 | 0.139 |
| Residence: Nomadic vs urban | -0.865 | 0.294 | 0.421 | 0.236–0.753 | 0.004 |
| Region: Marodijeh vs Awdal | 0.627 | 0.160 | 1.872 | 1.364–2.569 | <0.001 |
| Region: Sahil vs Awdal | 0.546 | 0.158 | 1.726 | 1.264–2.356 | 0.001 |
| Region: Togdheer vs Awdal | 0.611 | 0.157 | 1.842 | 1.351–2.510 | <0.001 |
| Region: Sool vs Awdal | 0.599 | 0.165 | 1.821 | 1.316–2.520 | <0.001 |
| Region: Sanaag vs Awdal | 0.123 | 0.168 | 1.131 | 0.812–1.576 | 0.465 |
| Constant | -3.419 | 0.320 | 0.033 | 0.017–0.062 | <0.001 |

N=14,399; overall model F(18,153)=28.05, p<0.001; global wealth Wald F(4,167)=0.63, p=0.6429; survey-model GOF F(9,162)=0.64, p=0.7655.

Note: Models 2 and 3 are alternative specifications relative to Model 1 rather than strictly sequential additions. All four models report the full retained regression results. β and SE are on the log-odds scale; where retained executed output provided OR and linearized SEs, β and SE were reconstructed as ln(OR) and SE(OR)/OR.

**Table S4. Adjusted predicted probabilities of any reported disability by household wealth quintile**

| **Wealth quintile** | **Adjusted probability** | **95% CI** |
| --- | --- | --- |
| Lowest | 7.04% | 4.21–9.87% |
| Second | 8.22% | 5.97–10.48% |
| Middle | 8.08% | 6.55–9.60% |
| Fourth | 7.34% | 5.75–8.93% |
| Highest | 6.86% | 5.37–8.34% |

Note: Predicted probabilities are from the primary multivariable model including wealth, categorical age group, sex, education, residence and region (N=14,399). Global wealth test: F(4,167)=0.63, p=0.6429. Pairwise wealth contrasts were additionally assessed with Bonferroni adjustment; no statistically detectable wealth contrast was identified.

**Table S5. Survey-weighted logistic regression for reported mental/psychological illness under alternative adjustment sets**

**Model 1: wealth + age + sex**

| **Term** | **β (log odds)** | **SE** | **OR** | **95% CI** | **P** |
| --- | --- | --- | --- | --- | --- |
| Wealth: Second vs lowest | 0.904 | 0.289 | 2.47 | 1.40–4.35 | 0.002 |
| Wealth: Middle vs lowest | 0.986 | 0.365 | 2.68 | 1.31–5.48 | 0.007 |
| Wealth: Fourth vs lowest | 0.489 | 0.511 | 1.63 | 0.60–4.45 | 0.336 |
| Wealth: Highest vs lowest | 0.122 | 0.612 | 1.13 | 0.34–3.74 | 0.836 |
| Age: 30–44 vs 18–29 | 0.247 | 0.329 | 1.28 | 0.67–2.43 | 0.453 |
| Age: 45–59 vs 18–29 | -0.186 | 0.431 | 0.83 | 0.36–1.95 | 0.674 |
| Age: ≥60 vs 18–29 | 0.507 | 0.419 | 1.66 | 0.73–3.77 | 0.224 |
| Sex: Female vs male | -0.528 | 0.223 | 0.59 | 0.38–0.91 | 0.018 |

N=14,579; 99 outcome events; overall model F(8,163)=4.87, p<0.001; global wealth Wald F(4,167)=3.36, p=0.0113; survey-model GOF F(9,162)=1.06, p=0.3970.

**Model 2: + education**

| **Term** | **β (log odds)** | **SE** | **OR** | **95% CI** | **P** |
| --- | --- | --- | --- | --- | --- |
| Wealth: Second vs lowest | 0.944 | 0.300 | 2.57 | 1.43–4.64 | 0.002 |
| Wealth: Middle vs lowest | 1.047 | 0.377 | 2.85 | 1.36–5.96 | 0.006 |
| Wealth: Fourth vs lowest | 0.615 | 0.455 | 1.85 | 0.76–4.53 | 0.177 |
| Wealth: Highest vs lowest | 0.351 | 0.747 | 1.42 | 0.33–6.17 | 0.640 |
| Age: 30–44 vs 18–29 | 0.131 | 0.357 | 1.14 | 0.57–2.31 | 0.704 |
| Age: 45–59 vs 18–29 | -0.274 | 0.441 | 0.76 | 0.32–1.80 | 0.528 |
| Age: ≥60 vs 18–29 | 0.365 | 0.481 | 1.44 | 0.56–3.69 | 0.448 |
| Sex: Female vs male | -0.654 | 0.263 | 0.52 | 0.31–0.87 | 0.014 |
| Education: Primary vs none | 0.223 | 0.384 | 1.25 | 0.59–2.66 | 0.554 |
| Education: Secondary vs none | -1.661 | 0.694 | 0.19 | 0.05–0.76 | 0.020 |
| Education: Higher vs none | -0.400 | 1.143 | 0.67 | 0.07–6.18 | 0.726 |

N=14,399; overall model F(11,160)=5.86, p<0.001; global wealth Wald F(4,167)=3.30, p=0.0124; survey-model GOF F(9,162)=2.05, p=0.0369.

**Model 3: + residence + region**

| **Term** | **β (log odds)** | **SE** | **OR** | **95% CI** | **P** |
| --- | --- | --- | --- | --- | --- |
| Wealth: Second vs lowest | 0.392 | 0.318 | 1.48 | 0.79–2.75 | 0.217 |
| Wealth: Middle vs lowest | 0.329 | 0.402 | 1.39 | 0.63–3.04 | 0.414 |
| Wealth: Fourth vs lowest | -0.151 | 0.509 | 0.86 | 0.32–2.35 | 0.771 |
| Wealth: Highest vs lowest | -0.446 | 0.748 | 0.64 | 0.15–2.81 | 0.552 |
| Age: 30–44 vs 18–29 | 0.131 | 0.356 | 1.14 | 0.57–2.30 | 0.704 |
| Age: 45–59 vs 18–29 | -0.274 | 0.457 | 0.76 | 0.31–1.86 | 0.552 |
| Age: ≥60 vs 18–29 | 0.372 | 0.476 | 1.45 | 0.57–3.69 | 0.430 |
| Sex: Female vs male | -0.673 | 0.266 | 0.51 | 0.30–0.85 | 0.010 |
| Education: Primary vs none | 0.174 | 0.393 | 1.19 | 0.55–2.57 | 0.649 |
| Education: Secondary vs none | -1.715 | 0.684 | 0.18 | 0.05–0.73 | 0.016 |
| Education: Higher vs none | -0.386 | 1.148 | 0.68 | 0.07–6.31 | 0.733 |
| Residence: Rural vs urban | -0.400 | 0.291 | 0.67 | 0.38–1.19 | 0.168 |
| Residence: Nomadic vs urban | -1.079 | 0.417 | 0.34 | 0.15–0.77 | 0.010 |
| Region: Marodijeh vs Awdal | 1.044 | 0.728 | 2.84 | 0.68–11.81 | 0.149 |
| Region: Sahil vs Awdal | 1.141 | 0.653 | 3.13 | 0.87–11.25 | 0.079 |
| Region: Togdheer vs Awdal | 1.275 | 0.642 | 3.58 | 1.02–12.61 | 0.047 |
| Region: Sool vs Awdal | 1.200 | 0.666 | 3.32 | 0.90–12.25 | 0.071 |
| Region: Sanaag vs Awdal | 0.673 | 0.659 | 1.96 | 0.54–7.16 | 0.307 |

Note: N=14,399; overall model F(18,153)=5.10, p<0.001; global wealth Wald F(4,167)=1.09, p=0.3655; Survey-model GOF F(9,162)=2.71, p=0.0058.

**Table S6. Survey-weighted logistic regression for concurrent reported disability and reported mental/psychological illness**

**Model 1: wealth + age + sex**

| **Term** | **β (log odds)** | **SE** | **OR** | **95% CI** | **P** |
| --- | --- | --- | --- | --- | --- |
| Wealth: Second vs lowest | 1.008 | 0.388 | 2.74 | 1.28–5.85 | 0.010 |
| Wealth: Middle vs lowest | 1.112 | 0.471 | 3.04 | 1.21–7.66 | 0.019 |
| Wealth: Fourth vs lowest | 0.507 | 0.470 | 1.66 | 0.66–4.16 | 0.281 |
| Wealth: Highest vs lowest | 0.405 | 0.740 | 1.50 | 0.35–6.36 | 0.580 |
| Age: 30–44 vs 18–29 | 0.322 | 0.376 | 1.38 | 0.66–2.88 | 0.390 |
| Age: 45–59 vs 18–29 | -0.186 | 0.569 | 0.83 | 0.27–2.51 | 0.741 |
| Age: ≥60 vs 18–29 | 0.344 | 0.425 | 1.41 | 0.61–3.23 | 0.420 |
| Sex: Female vs male | -0.545 | 0.303 | 0.58 | 0.32–1.05 | 0.072 |

Note: N=14,579; 68 co-occurrence events; overall model F(8,163)=3.01, p=0.0035; global wealth Wald F(4,167)=2.23, p=0.0674.

**Model 2: + education**

| **Term** | **β (log odds)** | **SE** | **OR** | **95% CI** | **P** |
| --- | --- | --- | --- | --- | --- |
| Wealth: Second vs lowest | 0.993 | 0.401 | 2.70 | 1.23–5.92 | 0.013 |
| Wealth: Middle vs lowest | 1.128 | 0.491 | 3.09 | 1.18–8.08 | 0.022 |
| Wealth: Fourth vs lowest | 0.571 | 0.418 | 1.77 | 0.78–4.02 | 0.174 |
| Wealth: Highest vs lowest | 0.588 | 0.911 | 1.80 | 0.30–10.68 | 0.517 |
| Age: 30–44 vs 18–29 | 0.239 | 0.419 | 1.27 | 0.56–2.89 | 0.568 |
| Age: 45–59 vs 18–29 | -0.236 | 0.583 | 0.79 | 0.25–2.46 | 0.680 |
| Age: ≥60 vs 18–29 | 0.285 | 0.459 | 1.33 | 0.54–3.26 | 0.530 |
| Sex: Female vs male | -0.654 | 0.314 | 0.52 | 0.28–0.96 | 0.038 |
| Education: Primary vs none | 0.615 | 0.336 | 1.85 | 0.96–3.59 | 0.067 |
| Education: Secondary vs none | -1.897 | 1.071 | 0.15 | 0.02–1.33 | 0.088 |
| Education: Higher vs none | -0.416 | 1.513 | 0.66 | 0.03–11.31 | 0.774 |

Note: N=14,399; overall model F(11,160)=4.37, p<0.001; global wealth Wald F(4,167)=2.00, p=0.0970.

**Model 3: + residence + region**

| **Term** | **β (log odds)** | **SE** | **OR** | **95% CI** | **P** |
| --- | --- | --- | --- | --- | --- |
| Wealth: Second vs lowest | 0.385 | 0.409 | 1.47 | 0.66–3.28 | 0.342 |
| Wealth: Middle vs lowest | 0.399 | 0.518 | 1.49 | 0.54–4.11 | 0.435 |
| Wealth: Fourth vs lowest | -0.236 | 0.551 | 0.79 | 0.27–2.34 | 0.674 |
| Wealth: Highest vs lowest | -0.288 | 0.888 | 0.75 | 0.13–4.23 | 0.746 |
| Age: 30–44 vs 18–29 | 0.215 | 0.423 | 1.24 | 0.54–2.84 | 0.605 |
| Age: 45–59 vs 18–29 | -0.261 | 0.595 | 0.77 | 0.24–2.47 | 0.661 |
| Age: ≥60 vs 18–29 | 0.262 | 0.464 | 1.30 | 0.52–3.21 | 0.571 |
| Sex: Female vs male | -0.693 | 0.316 | 0.50 | 0.27–0.93 | 0.030 |
| Education: Primary vs none | 0.536 | 0.348 | 1.71 | 0.86–3.36 | 0.123 |
| Education: Secondary vs none | -1.966 | 1.042 | 0.14 | 0.02–1.19 | 0.072 |
| Education: Higher vs none | -0.478 | 1.512 | 0.62 | 0.03–11.24 | 0.748 |
| Residence: Rural vs urban | -0.274 | 0.409 | 0.76 | 0.34–1.69 | 0.497 |
| Residence: Nomadic vs urban | -1.238 | 0.546 | 0.29 | 0.10–0.85 | 0.023 |
| Region: Marodijeh vs Awdal | 0.837 | 0.748 | 2.31 | 0.53–9.95 | 0.261 |
| Region: Sahil vs Awdal | 0.713 | 0.671 | 2.04 | 0.55–7.64 | 0.286 |
| Region: Togdheer vs Awdal | 0.798 | 0.670 | 2.22 | 0.60–8.30 | 0.233 |
| Region: Sool vs Awdal | 0.708 | 0.697 | 2.03 | 0.52–7.99 | 0.307 |
| Region: Sanaag vs Awdal | 0.166 | 0.682 | 1.18 | 0.31–4.50 | 0.805 |

Note: N=14,399; overall model F(18,153)=3.37, p<0.001; global wealth Wald F(4,167)=0.72, p=0.5787.

Note: Reference categories are shown in term labels. Because 68 co-occurrence events were observed, secondary regression estimates are sparse and should be interpreted cautiously. Survey-model GOF statistics were F(9,162)=0.55, p=0.8327 for Model 1; F(9,162)=450.58, p<0.001 for Model 2; and F(9,162)=700.51, p<0.001 for Model 3.

**Table S7. Concentration indices for socioeconomic inequality in reported disability, reported mental/psychological illness, and co-occurrence**

| Outcome | Concentration index | SE | 95% CI | P value | Erreygers index | Erreygers SE / 95% CI / P value |
| --- | --- | --- | --- | --- | --- | --- |
| Any reported disability | 0.0584 | 0.0284 | 0.0023–0.1144 | 0.0415 | 0.0171 | 0.0083 / 0.0007–0.0334 / 0.0415 |
| Reported mental/psychological illness | -0.0703 | 0.0685 | -0.2056–0.0650 | 0.3063 | -0.0020 | 0.0019 / -0.0058–0.0018 / 0.3063 |
| Disability + reported mental/psychological illness | -0.0432 | 0.0901 | -0.2210–0.1346 | 0.6323 | -0.0009 | 0.0018 / -0.0044–0.0027 / 0.6323 |

Note: Wealth ranking used the continuous household wealth-index factor score, ordered from poorest to richest. Positive values indicate relative concentration toward higher wealth ranks; negative values indicate concentration toward lower ranks. Concentration-index estimates are reported with SEs and 95% CIs; the Erreygers index is reported with its SE, 95% CI and P value in the final column. These are marginal distributional measures and answer a different question from adjusted regression models.

**Table S8. Domain-specific concentration indices for the seven reported disability domains**

| **Disability domain** | **Concentration index** | **SE** | **95% CI** | **P** |
| --- | --- | --- | --- | --- |
| Sight | 0.0504 | 0.0521 | -0.0524–0.1532 | 0.3343 |
| Hearing | -0.0216 | 0.0636 | -0.1472–0.1040 | 0.7344 |
| Speech | -0.0320 | 0.1054 | -0.2401–0.1760 | 0.7616 |
| Learning | 0.1155 | 0.1481 | -0.1768–0.4078 | 0.4363 |
| Mobility | 0.0372 | 0.0632 | -0.0875–0.1619 | 0.5566 |
| Self-care | -0.2408 | 0.0849 | -0.4084–-0.0732 | 0.0051 |
| Mental disability domain | 0.1106 | 0.0622 | -0.0123–0.2335 | 0.0773 |

Note: Concentration indices used the continuous household wealth-index factor score as the ranking variable. Estimates are reported with survey-based SEs, 95% CIs and P values. The self-care result was statistically detectable but should be interpreted cautiously because the domain is uncommon.

**Table S9. Assessment of missing education data**

| **Measure** | **Complete education data** | **Education missing** |
| --- | --- | --- |
| Number of observations | 14,399 | 180 |
| Percentage of sample | 98.77% | 1.23% |
| Weighted any-disability prevalence | 7.29% | 8.56% |
| Weighted mental/psychological illness prevalence | 0.72% | Not reported (0 observed cases) |
| Weighted co-occurrence prevalence | 0.51% | Not reported (0 observed cases) |
| Association of education missingness with any disability | Reference | OR 1.19 (95% CI 0.49–2.88), p=0.698 |

Note: The missingness analysis does not establish that education data were missing completely at random. Complete-case estimates were retained for models requiring education. Logistic model: education missingness regressed on any reported disability, N=14,579, OR 1.19, 95% CI 0.49–2.88, p=0.698. β and SE can be represented as −0.169 and 0.453 on the log-odds scale. Overall model test: F(1,170)=0.15, p=0.6982. Survey-model GOF F(9,162)=0.00, p=1.0000.

**Table S10. Sensitivity analysis using continuous age instead of categorical age**

| **Specification** | **Age effect** | **β (log odds)** | **SE** | **95% CI** | **P value** | **Comment** |
| --- | --- | --- | --- | --- | --- | --- |
| Primary model (categorical age) | 45–59 vs 18–29: OR 1.56; ≥60 vs 18–29: OR 6.95 | See primary model | See primary model | 1.05–2.32; 5.05–9.56 | 0.029; <0.001 | Joint age Wald F(3,168)=74.51, p<0.001 |
| Sensitivity model (continuous age) | OR 1.042 per additional year | 0.042 | 0.003 | 1.035–1.049 | <0.001 | Categorical age retained for primary model to avoid imposing linearity across the full age range. |

Note: The continuous-age sensitivity model retains wealth, sex, education, residence and region and replaces age group with continuous age. The continuous-age β and SE are taken from the supplied Stata OR and linearized standard error. Overall model test: F(16,155)=26.81, p<0.001. Survey-model GOF F(9,162)=2.74, p=0.0053.

**Table S11. Sparse-event distribution for reported mental/psychological illness and disability–mental/psychological illness co-occurrence**

| **Predictor** | **Category** | **Mental illness cases n** | **Co-occurrence cases n** | **Total N** |
| --- | --- | --- | --- | --- |
| Wealth | Lowest | 29 | 18 | 6,146 |
| Wealth | Second | 19 | 14 | 1,521 |
| Wealth | Middle | 19 | 13 | 1,839 |
| Wealth | Fourth | 21 | 14 | 2,481 |
| Wealth | Highest | 11 | 9 | 2,592 |
| Age group | 18–29 | 38 | 25 | 5,753 |
| Age group | 30–44 | 27 | 17 | 4,396 |
| Age group | 45–59 | 14 | 11 | 2,378 |
| Age group | ≥60 | 20 | 15 | 2,052 |
| Sex | Male | 56 | 37 | 6,708 |
| Sex | Female | 43 | 31 | 7,871 |
| Education | No education | 81 | 55 | 10,892 |
| Education | Primary | 12 | 11 | 1,588 |
| Education | Secondary | 4 | 1 | 1,199 |
| Education | Higher | 2 | 1 | 720 |
| Education | Missing | 0 | 0 | 180 |
| Residence | Urban | 46 | 33 | 5,381 |
| Residence | Rural | 34 | 25 | 4,470 |
| Residence | Nomadic | 19 | 10 | 4,728 |
| Region | Awdal | 5 | 5 | 2,196 |
| Region | Marodijeh | 17 | 14 | 2,036 |
| Region | Sahil | 15 | 10 | 1,793 |
| Region | Togdheer | 22 | 15 | 2,442 |
| Region | Sool | 25 | 15 | 2,971 |
| Region | Sanaag | 15 | 9 | 3,141 |

Note: Counts are unweighted and correspond to the full analytic dataset (N=14,579). Mental/psychological illness had 99 total events and co-occurrence had 68 events. Sparse cells, particularly in education categories, limit precision of detailed models.

**Table S12. Survey-weighted prevalence of the seven disability domains by sex**

| **Disability domain** | **Male % (95% CI)** | **Female % (95% CI)** | **P value** |
| --- | --- | --- | --- |
| Sight | 3.46 (2.66–4.49) | 4.14 (3.44–4.97) | 0.1506 |
| Hearing | 1.03 (0.64–1.66) | 1.95 (1.50–2.53) | 0.0129 |
| Speech | 0.25 (0.15–0.44) | 0.44 (0.24–0.83) | 0.2095 |
| Learning | 0.42 (0.19–0.93) | 0.29 (0.16–0.52) | 0.4934 |
| Mobility | 1.38 (1.07–1.77) | 1.40 (1.07–1.83) | 0.9300 |
| Self-care | 0.28 (0.18–0.45) | 0.30 (0.20–0.46) | 0.8463 |
| Mental disability domain | 1.54 (1.13–2.09) | 1.08 (0.75–1.55) | 0.0729 |

Note: Values are survey-weighted prevalence estimates with 95% confidence intervals. P values are from survey-adjusted design-based comparisons. Domain indicators are not mutually exclusive.

**Table S13. Survey-weighted prevalence of the seven disability domains by adult age group**

| **Disability domain** | **18–29 % (95% CI)** | **30–44 % (95% CI)** | **45–59 % (95% CI)** | **60+ % (95% CI)** | **P value** |
| --- | --- | --- | --- | --- | --- |
| Sight | 1.73 (1.35–2.22) | 1.79 (1.15–2.77) | 3.98 (2.78–5.67) | 14.72 (12.08–17.82) | <0.001 |
| Hearing | 0.81 (0.49–1.32) | 0.69 (0.43–1.12) | 0.91 (0.49–1.70) | 6.38 (4.48–9.02) | <0.001 |
| Speech | 0.24 (0.11–0.52) | 0.35 (0.10–1.27) | 0.23 (0.08–0.67) | 0.89 (0.44–1.79) | 0.0873 |
| Learning | 0.25 (0.11–0.58) | 0.27 (0.05–1.44) | 0.11 (0.03–0.42) | 1.12 (0.67–1.86) | 0.0012 |
| Mobility | 0.86 (0.60–1.23) | 0.65 (0.46–0.92) | 1.20 (0.80–1.81) | 4.90 (3.76–6.35) | <0.001 |
| Self-care | 0.17 (0.09–0.30) | 0.25 (0.15–0.41) | 0.27 (0.12–0.60) | 0.81 (0.51–1.27) | <0.001 |
| Mental disability domain | 1.28 (0.82–2.00) | 1.36 (0.97–1.92) | 0.50 (0.26–0.98) | 2.03 (1.11–3.68) | 0.0107 |

Note: Values are survey-weighted prevalence estimates with 95% confidence intervals. P values are from survey-adjusted Wald tests across the four ordered age groups. Domain indicators are not mutually exclusive.

**Table S14. Ordered age-trend analysis for any reported disability**

| **Outcome** | **β (log odds)** | **SE** | **OR per one-category increase** | **95% CI** | **P value** | **Design-based test** |
| --- | --- | --- | --- | --- | --- | --- |
| Any reported disability | 0.683 | 0.055 | 1.98 | 1.78–2.21 | <0.001 | F(1,170)=153.68; p<0.001 |

Note: The four age groups (18–29, 30–44, 45–59, ≥60 years) were entered as an ordered categorical predictor. This tests an ordered gradient and does not assume equal year intervals between categories. Overall model test: F(1,170)=153.68, p<0.001. Survey-model GOF F(9,162)=4.78, p<0.001.

**Table S15. Survey-weighted joint distribution of reported disability and reported mental/psychological illness and assessment of departure from independence**

| **Measure** | **Estimate** | **95% CI / test** | **Interpretation** |
| --- | --- | --- | --- |
| Any reported disability | 7.3045% | 6.6191–7.9899% | Marginal prevalence |
| Reported mental/psychological illness | 0.7099% | 0.4640–0.9558% | Marginal prevalence |
| Both reported disability + reported mental/psychological illness | 0.5032% | 0.2832–0.7232% | Observed joint prevalence |
| Expected joint prevalence under independence | 0.05185% | 0.03195–0.07176% | Product of marginal prevalences |
| Observed − expected | 0.45134 percentage points | 0.24991–0.65276 points | Departure from independence |
| Observed/expected ratio | 9.70 | 7.60–11.81 | Observed joint prevalence relative to independence |
| Design-based Pearson test | F(1,170)=239.54 | p<0.001 | Formal test of association |

**Adjusted association: mental/psychological illness as outcome**

| **Term** | **β (log odds)** | **SE** | **OR** | **95% CI** | **P** |
| --- | --- | --- | --- | --- | --- |
| Any reported disability | 3.761 | 0.432 | 42.99 | 18.45–100.18 | <0.001 |

Note: Primary model adjusted for age group and sex; N=14,579; 99 mental/psychological illness events. Survey-model GOF F(9,162)=0.33, p=0.9623.

**Sensitivity adjusted association**

| **Term** | **β (log odds)** | **SE** | **OR** | **95% CI** | **P** |
| --- | --- | --- | --- | --- | --- |
| Any reported disability | 3.390 | 0.428 | 29.68 | 12.83–68.66 | <0.001 |

Note: Sensitivity model additionally adjusted for marital status and household size. Survey-model GOF F(9,162)=495778.97, p<0.001.

Note: Expected joint prevalence under independence equals the product of the two marginal weighted prevalences. The adjusted association models are cross-sectional and do not establish direction or causality. The displayed β and SE are derived from the reported OR and 95% CI because the supplied table did not contain the original coefficient/SE columns.

**Table S16. Survey-weighted four-category co-occurrence by education, residence and region**

| Dimension | Group | Neither % (n/N) | Disability only % (n/N) | Mental/psychological illness only % (n/N) | Both % (n/N) | Global design-based test |
| --- | --- | --- | --- | --- | --- | --- |
| Education | No education | 91.32 (10,064/10,892) | 7.91 (747/10,892) | 0.27 (26/10,892) | 0.50 (55/10,892) | F(5.08,863.01)=3.74; p=0.0022 |
|  | Primary | 93.89 (1,511/1,588) | 5.07 (65/1,588) | 0.02 (1/1,588) | 1.03 (11/1,588) |  |
|  | Secondary | 95.80 (1,163/1,199) | 4.04 (32/1,199) | 0.07 (3/1,199) | 0.09 (1/1,199) |  |
|  | Higher | 95.22 (692/720) | 4.21 (26/720) | 0.16 (1/720) | 0.40 (1/720) |  |
| Residence | Urban | 91.43 (4,961/5,381) | 7.76 (374/5,381) | 0.22 (13/5,381) | 0.58 (33/5,381) | F(4.83,821.49)=8.90; p<0.001 |
|  | Rural | 91.90 (4,096/4,470) | 7.34 (340/4,470) | 0.18 (9/4,470) | 0.58 (25/4,470) |  |
|  | Nomadic | 96.24 (4,544/4,728) | 3.39 (165/4,728) | 0.19 (9/4,728) | 0.18 (10/4,728) |  |
| Region | Awdal | 95.04 (2,092/2,196) | 4.69 (99/2,196) | 0.00† (0/2,196) | 0.27 (5/2,196) | F(8.80,1495.53)=2.80; p=0.0031 |
|  | Marodijeh | 91.33 (1,875/2,036) | 7.96 (144/2,036) | 0.12 (3/2,036) | 0.58 (14/2,036) |  |
|  | Sahil | 92.84 (1,656/1,793) | 6.38 (122/1,793) | 0.27 (5/1,793) | 0.50 (10/1,793) |  |
|  | Togdheer | 91.40 (2,251/2,442) | 7.57 (169/2,442) | 0.38 (7/2,442) | 0.65 (15/2,442) |  |
|  | Sool | 92.52 (2,746/2,971) | 6.66 (200/2,971) | 0.34 (10/2,971) | 0.49 (15/2,971) |  |
|  | Sanaag | 95.32 (2,981/3,141) | 4.25 (145/3,141) | 0.19 (6/3,141) | 0.25 (9/3,141) |  |

Note: Values are survey-weighted prevalence within each subgroup; unweighted numerators and denominators are shown in parentheses. Education estimates exclude 180 respondents with missing education data (N=14,399 for the education comparison); residence and region estimates use the full analytic sample (N=14,579). Global design-based P values are from Pearson tests of the four-category distribution. † No observed mental/psychological illness-only cases occurred in Awdal; this is an observed sample cell of zero and should not be interpreted as evidence that population prevalence is zero.

**Table S17. Reported origin of disability among adults with reported disability**

| **Reported main origin** | Weighted % (n/N) | **95% CI** |
| --- | --- | --- |
| Congenital | 10.02 (73/799) | 7.20–13.79 |
| Contagious disease | 10.71 (100/799) | 7.93–14.31 |
| Child birth conditions | 2.37 (29/799) | 1.42–3.92 |
| Other disease | 23.81 (152/799) | 19.70–28.47 |
| Abuse | 0.20 (4/799) | 0.06–0.70 |
| Aging | 30.93 (262/799) | 26.57–35.65 |
| Injury/accident | 9.96 (93/799) | 7.63–12.90 |
| Witchcraft | 0.13 (2/799) | 0.03–0.54 |
| Others | 4.47 (34/799) | 2.44–8.03 |
| Don't know | 7.40 (50/799) | 5.23–10.37 |

Note: Among 799 adults with reported disability and valid origin responses. Values are survey-weighted prevalence estimates; unweighted numerators and denominators are shown in parentheses. These are respondent-reported contextual attributions and are not causal fractions.

**Table S18. Reported age at onset of disability among adults with valid onset data**

| **Reported age at onset** | Weighted % (n/N) | **95% CI** |
| --- | --- | --- |
| <5 years | 15.59 (126/799) | 12.08–19.90 |
| 5–9 years | 5.95 (44/799) | 3.75–9.31 |
| 10–19 years | 11.06 (75/799) | 8.00–15.11 |
| 20–29 years | 11.07 (79/799) | 6.69–17.75 |
| 30–39 years | 6.63 (54/799) | 4.44–9.79 |
| 40–49 years | 12.71 (87/799) | 9.04–17.59 |
| 50–59 years | 10.33 (99/799) | 7.91–13.39 |
| 60–69 years | 13.16 (119/799) | 10.52–16.35 |
| 70+ years | 13.50 (116/799) | 9.87–18.18 |

Note: Among 799 adults with reported disability and valid age-at-onset information. Values are survey-weighted prevalence estimates; unweighted numerators and denominators are shown in parentheses. Categories reproduce the source-consistent grouping used in the SLHDS disability table. These are reported timing categories and are not incidence estimates.

**Table S19. Reported care/support among adults with reported disability**

| **Support category** | Weighted % (n/N) | **95% CI** |
| --- | --- | --- |
| Medical support | 79.95 (770/921) | 76.42–83.07 |
| Welfare support | 0.56 (10/921) | 0.20–1.54 |
| Financial support | 2.56 (33/921) | 1.60–4.09 |
| Nutritional support | 0.96 (6/921) | 0.31–2.93 |
| No support | 29.86 (234/921) | 27.22–32.65 |

Note: Among 921 adults with reported disability and at least one recorded support indicator. Values are survey-weighted prevalence estimates; unweighted numerators and denominators are shown in parentheses. Support categories are multiple-response indicators, so percentages are calculated separately and do not sum to 100%.

**Table S20. Sensitivity analysis excluding records with both “None” and a positive disability domain**

| **Measure** | **Primary analysis** | **Excluding 227 conflict records** |
| --- | --- | --- |
| Analytic observations | 14,579 | 14,352 |
| Weighted prevalence of any reported disability | 7.30% (947/14,579) (6.65–8.02%) | 5.13% (720/14,352) (4.58–5.73%) |
| Complete-case adjusted model N | 14,399 | 14,174 |
| Global wealth Wald test | F(4,167)=0.63; p=0.6429 | F(4,167)=0.86; p=0.4909 |

**Excluding the 227 conflict records: adjusted wealth terms**

| Term | β (log odds) | SE | OR | 95% CI | P |
| --- | --- | --- | --- | --- | --- |
| Wealth: Second vs lowest | 0.223 | 0.295 | 1.250 | 0.698–2.239 | 0.451 |
| Wealth: Middle vs lowest | 0.121 | 0.291 | 1.129 | 0.636–2.006 | 0.677 |
| Wealth: Fourth vs lowest | -0.068 | 0.327 | 0.935 | 0.490–1.782 | 0.836 |
| Wealth: Highest vs lowest | -0.157 | 0.340 | 0.855 | 0.437–1.672 | 0.644 |
| Age: 30–44 vs 18–29 | 0.649 | 0.265 | 1.913 | 1.133–3.228 | 0.015 |
| Age: 45–59 vs 18–29 | 1.293 | 0.272 | 3.645 | 2.130–6.238 | <0.001 |
| Age: ≥60 vs 18–29 | 2.684 | 0.251 | 14.645 | 8.916–24.055 | <0.001 |
| Female vs male | 0.107 | 0.132 | 1.113 | 0.859–1.443 | 0.416 |
| Education: Primary vs none | 0.247 | 0.207 | 1.280 | 0.850–1.927 | 0.236 |
| Education: Secondary vs none | -0.016 | 0.228 | 0.984 | 0.627–1.543 | 0.943 |
| Education: Higher vs none | -0.587 | 0.430 | 0.556 | 0.238–1.299 | 0.174 |
| Residence: Rural vs urban | -0.179 | 0.155 | 0.836 | 0.616–1.135 | 0.248 |
| Residence: Nomadic vs urban | -0.513 | 0.307 | 0.599 | 0.326–1.099 | 0.097 |
| Region: Marodijeh vs Awdal | 0.589 | 0.184 | 1.802 | 1.254–2.590 | 0.002 |
| Region: Sahil vs Awdal | 0.689 | 0.168 | 1.991 | 1.430–2.773 | <0.001 |
| Region: Togdheer vs Awdal | 0.740 | 0.162 | 2.095 | 1.523–2.883 | <0.001 |
| Region: Sool vs Awdal | 0.696 | 0.179 | 2.006 | 1.409–2.854 | <0.001 |
| Region: Sanaag vs Awdal | 0.283 | 0.180 | 1.328 | 0.931–1.893 | 0.117 |
| Constant | -4.533 | 0.390 | 0.011 | 0.005–0.023 | <0.001 |

Note: N=14,174; overall model F(18,153)=23.94, p<0.001; global wealth Wald F(4,167)=0.86, p=0.4909. Survey-model GOF was F(9,162)=0.87, p=0.5525.

Note: The primary disability definition uses the seven labelled positive domain indicators as authoritative. A source-data inconsistency was identified in 227 adults in whom the “None” indicator co-occurred with at least one positive disability-domain indicator. The sensitivity analysis excluded these 227 records and repeated the primary survey-weighted disability prevalence estimate and adjusted regression model. Values for prevalence are survey-weighted estimates with unweighted numerators and denominators shown in parentheses. The adjusted model included wealth quintile, age group, sex, education, residence, and region and used complete cases (N=14,174). Reference categories were the lowest wealth quintile, age 18–29 years, male sex, no education, urban residence, and Awdal region. All estimates account for the complex survey design. β and SE are on the log-odds scale; where the retained output provided OR and its linearized SE, β and SE are reconstructed as ln(OR) and SE(OR)/OR.

**Table S21. Covariate-specification sensitivity models for reported disability**

**Primary model: + age group, sex, education, residence, region**

| Term | β (log odds) | SE | OR | 95% CI | P |
| --- | --- | --- | --- | --- | --- |
| Wealth: Second vs lowest | 0.183 | 0.268 | 1.200 | 0.707–2.039 | 0.497 |
| Wealth: Middle vs lowest | 0.162 | 0.282 | 1.175 | 0.673–2.052 | 0.568 |
| Wealth: Fourth vs lowest | 0.049 | 0.296 | 1.050 | 0.586–1.883 | 0.869 |
| Wealth: Highest vs lowest | -0.031 | 0.311 | 0.970 | 0.525–1.790 | 0.922 |
| Age: 30–44 vs 18–29 | 0.066 | 0.165 | 1.068 | 0.772–1.478 | 0.688 |
| Age: 45–59 vs 18–29 | 0.444 | 0.202 | 1.559 | 1.047–2.321 | 0.029 |
| Age: ≥60 vs 18–29 | 1.939 | 0.162 | 6.949 | 5.051–9.561 | <0.001 |
| Female vs male | -0.001 | 0.101 | 0.999 | 0.819–1.219 | 0.994 |
| Education: Primary vs none | 0.017 | 0.159 | 1.017 | 0.743–1.392 | 0.916 |
| Education: Secondary vs none | -0.405 | 0.234 | 0.667 | 0.420–1.059 | 0.085 |
| Education: Higher vs none | -0.350 | 0.300 | 0.705 | 0.390–1.274 | 0.245 |
| Residence: Rural vs urban | -0.218 | 0.146 | 0.804 | 0.602–1.074 | 0.139 |
| Residence: Nomadic vs urban | -0.865 | 0.294 | 0.421 | 0.236–0.753 | 0.004 |
| Region: Marodijeh vs Awdal | 0.627 | 0.160 | 1.872 | 1.364–2.569 | <0.001 |
| Region: Sahil vs Awdal | 0.546 | 0.158 | 1.726 | 1.264–2.356 | 0.001 |
| Region: Togdheer vs Awdal | 0.611 | 0.157 | 1.842 | 1.351–2.510 | <0.001 |
| Region: Sool vs Awdal | 0.599 | 0.165 | 1.821 | 1.316–2.520 | <0.001 |
| Region: Sanaag vs Awdal | 0.123 | 0.168 | 1.131 | 0.812–1.576 | 0.465 |
| Constant | -3.419 | 0.320 | 0.033 | 0.017–0.062 | <0.001 |

N=14,399; overall model F(18,153)=28.05, p<0.001; global wealth Wald F(4,167)=0.63, p=0.6429; survey-model GOF F(9,162)=0.64, p=0.7655.

**+ marital status + household size**

| Term | β (log odds) | SE | OR | 95% CI | P |
| --- | --- | --- | --- | --- | --- |
| Wealth: Second vs lowest | 0.160 | 0.272 | 1.173 | 0.686–2.006 | 0.557 |
| Wealth: Middle vs lowest | 0.232 | 0.289 | 1.262 | 0.713–2.232 | 0.422 |
| Wealth: Fourth vs lowest | 0.116 | 0.304 | 1.123 | 0.617–2.045 | 0.703 |
| Wealth: Highest vs lowest | 0.010 | 0.314 | 1.010 | 0.543–1.880 | 0.974 |
| Age: 30–44 vs 18–29 | 0.770 | 0.173 | 2.160 | 1.535–3.040 | <0.001 |
| Age: 45–59 vs 18–29 | 1.260 | 0.212 | 3.524 | 2.317–5.360 | <0.001 |
| Age: ≥60 vs 18–29 | 2.596 | 0.203 | 13.415 | 8.979–20.044 | <0.001 |
| Female vs male | -0.056 | 0.120 | 0.946 | 0.747–1.198 | 0.641 |
| Education: Primary vs none | 0.121 | 0.157 | 1.128 | 0.827–1.539 | 0.444 |
| Education: Secondary vs none | -0.449 | 0.234 | 0.638 | 0.402–1.013 | 0.056 |
| Education: Higher vs none | -0.380 | 0.290 | 0.684 | 0.386–1.211 | 0.191 |
| Residence: Rural vs urban | -0.147 | 0.158 | 0.863 | 0.632–1.178 | 0.351 |
| Residence: Nomadic vs urban | -0.728 | 0.307 | 0.483 | 0.264–0.884 | 0.019 |
| Region: Marodijeh vs Awdal | 0.640 | 0.168 | 1.896 | 1.360–2.643 | <0.001 |
| Region: Sahil vs Awdal | 0.564 | 0.161 | 1.757 | 1.280–2.413 | 0.001 |
| Region: Togdheer vs Awdal | 0.670 | 0.159 | 1.954 | 1.426–2.676 | <0.001 |
| Region: Sool vs Awdal | 0.643 | 0.166 | 1.901 | 1.371–2.637 | <0.001 |
| Region: Sanaag vs Awdal | 0.164 | 0.168 | 1.178 | 0.846–1.641 | 0.329 |
| Marital status: Divorced vs married | 0.258 | 0.209 | 1.294 | 0.857–1.953 | 0.218 |
| Marital status: Abandoned vs married | 0.539 | 0.496 | 1.715 | 0.645–4.561 | 0.278 |
| Marital status: Widowed vs married | 0.846 | 0.116 | 2.330 | 1.855–2.927 | <0.001 |
| Marital status: Never married vs married | 1.447 | 0.158 | 4.250 | 3.113–5.802 | <0.001 |
| Household size | -0.043 | 0.023 | 0.958 | 0.915–1.004 | 0.071 |
| Constant | -4.241 | 0.433 | 0.014 | 0.006–0.034 | <0.001 |

N=14,399; overall model F(23,148)=22.59, p<0.001; global wealth Wald F(4,167)=0.57, p=0.6874; survey-model GOF F(9,162)=5.83, p<0.001.

**+ marital status + household size + relationship to head**

| Term | β (log odds) | SE | OR | 95% CI | P |
| --- | --- | --- | --- | --- | --- |
| Wealth: Second vs lowest | 0.140 | 0.282 | 1.151 | 0.661–2.001 | 0.618 |
| Wealth: Middle vs lowest | 0.176 | 0.297 | 1.193 | 0.667–2.133 | 0.551 |
| Wealth: Fourth vs lowest | 0.013 | 0.312 | 1.013 | 0.549–1.868 | 0.967 |
| Wealth: Highest vs lowest | -0.116 | 0.326 | 0.891 | 0.468–1.695 | 0.722 |
| Age: 30–44 vs 18–29 | 0.790 | 0.179 | 2.203 | 1.548–3.136 | <0.001 |
| Age: 45–59 vs 18–29 | 1.274 | 0.217 | 3.576 | 2.329–5.490 | <0.001 |
| Age: ≥60 vs 18–29 | 2.495 | 0.206 | 12.123 | 8.070–18.210 | <0.001 |
| Female vs male | 0.017 | 0.122 | 1.017 | 0.800–1.294 | 0.890 |
| Education: Primary vs none | 0.200 | 0.165 | 1.222 | 0.883–1.691 | 0.225 |
| Education: Secondary vs none | -0.357 | 0.236 | 0.699 | 0.439–1.113 | 0.131 |
| Education: Higher vs none | -0.326 | 0.286 | 0.722 | 0.410–1.270 | 0.256 |
| Residence: Rural vs urban | -0.156 | 0.160 | 0.856 | 0.623–1.174 | 0.332 |
| Residence: Nomadic vs urban | -0.685 | 0.312 | 0.504 | 0.272–0.934 | 0.030 |
| Region: Marodijeh vs Awdal | 0.696 | 0.173 | 2.006 | 1.426–2.821 | <0.001 |
| Region: Sahil vs Awdal | 0.602 | 0.159 | 1.826 | 1.336–2.498 | <0.001 |
| Region: Togdheer vs Awdal | 0.687 | 0.163 | 1.987 | 1.442–2.739 | <0.001 |
| Region: Sool vs Awdal | 0.697 | 0.168 | 2.007 | 1.441–2.795 | <0.001 |
| Region: Sanaag vs Awdal | 0.201 | 0.168 | 1.222 | 0.877–1.702 | 0.234 |
| Marital status: Divorced vs married | -0.013 | 0.184 | 0.987 | 0.686–1.420 | 0.944 |
| Marital status: Abandoned vs married | 0.171 | 0.539 | 1.186 | 0.409–3.437 | 0.752 |
| Marital status: Widowed vs married | 0.559 | 0.123 | 1.749 | 1.372–2.230 | <0.001 |
| Marital status: Never married vs married | 0.915 | 0.239 | 2.497 | 1.557–4.005 | <0.001 |
| Household size | -0.060 | 0.024 | 0.941 | 0.897–0.988 | 0.014 |
| Relationship: Spouse vs head | -0.405 | 0.186 | 0.667 | 0.462–0.962 | 0.030 |
| Relationship: Son/Daughter vs head | 0.341 | 0.217 | 1.407 | 0.917–2.158 | 0.117 |
| Relationship: Son-in-law/Daughter-in-law vs head | 1.022 | 0.474 | 2.778 | 1.091–7.073 | 0.032 |
| Relationship: Grandchild vs head | 0.070 | 0.815 | 1.073 | 0.215–5.359 | 0.932 |
| Relationship: Parent vs head | 0.749 | 0.251 | 2.116 | 1.289–3.473 | 0.003 |
| Relationship: Parent-in-law vs head | 0.578 | 0.316 | 1.783 | 0.955–3.326 | 0.069 |
| Relationship: Brother/Sister vs head | 0.751 | 0.305 | 2.120 | 1.161–3.872 | 0.015 |
| Relationship: Nephew/Niece vs head | 0.845 | 0.325 | 2.328 | 1.225–4.424 | 0.010 |
| Relationship: Brother/Sister-in-law vs head | 1.068 | 0.355 | 2.910 | 1.445–5.860 | 0.003 |
| Relationship: Other Relative vs head | 0.612 | 0.300 | 1.843 | 1.020–3.332 | 0.043 |
| Relationship: Adopted/Foster/Step Child vs head | 1.708 | 0.542 | 5.516 | 1.892–16.086 | 0.002 |
| Relationship: Not Related vs head | -0.332 | 0.967 | 0.718 | 0.106–4.841 | 0.732 |
| Relationship: Don't Know vs head | 2.154 | 0.989 | 8.615 | 1.224–60.641 | 0.031 |
| Constant | -4.126 | 0.438 | 0.016 | 0.007–0.038 | <0.001 |

N=14,399; overall model F(36,135)=31.49, p<0.001; global wealth Wald F(4,167)=0.85, p=0.4942; survey-model GOF F(9,162)=4.53, p<0.001.

Note: All specifications use the complete-education analytic sample (N=14,399) and incorporate the complex survey design. The primary model is defined as adjustment for wealth quintile, age group, sex, education, residence, and region; sensitivity specification 1 additionally includes marital status and household size, and sensitivity specification 2 further includes relationship to household head. Reference categories are the omitted categories in the Stata specifications: lowest wealth quintile, age 18–29 years, male sex, no education, urban residence, Awdal region, married marital status, and household head. β coefficients and SEs are on the log-odds scale; ORs are adjusted odds ratios with 95% confidence intervals. Where retained executed output provided ORs and linearized SEs rather than the original coefficient table, β and SE were reconstructed as ln(OR) and SE(OR)/OR. Relationship-to-household-head categories are retained for completeness; several categories have wide confidence intervals, reflecting limited precision.
